# Impact of unequal testing on vaccine effectiveness estimates across two study designs: a simulation study

**DOI:** 10.1101/2024.08.27.24312655

**Authors:** Korryn Bodner, Linwei Wang, Rafal Kustra, Jeffrey C. Kwong, Beate Sander, Hind Sbihi, Michael A Irvine, Sharmistha Mishra

**Affiliations:** MAP Centre for Urban Health Solutions, Li Ka Shing Knowledge Institute, St. Michael’s Hospital, Unity Health Toronto, Toronto, Canada; Division of Biostatistics, Dalla Lana School of Public Health, University of Toronto, Toronto, ON, Canada; Department of Statistical Sciences, University of Toronto, Toronto, ON, Canada; ICES, Toronto, Canada; Dalla Lana School of Public Health, University of Toronto, Toronto, Canada; Public Health Ontario, Toronto, ON, Canada; Department of Family and Community Medicine, University of Toronto, Toronto, ON, Canada; University Health Network, Toronto, ON, Canada; Institute of Health Policy, Management and Evaluation, Dalla Lana School of Public Health, University of Toronto, Toronto, Canada; Toronto Health Economics and Technology Assessment (THETA) Collaborative, University Health Network, Toronto, Canada; British Columbia Centre for Disease Control, Vancouver, Canada; University of British Columbia, School of Population and Public Health, Vancouver, BC, Canada; Faculty of Health Sciences, Simon Fraser University, Burnaby, Canada; Institute of Medical Science, University of Toronto, Toronto, Canada; Division of Infectious Diseases, Department of Medicine, University of Toronto, Toronto, Canada

**Author notes:** **Corresponding Author:** Dr. Korryn Bodner, 326.82, 3rd Floor, St. Michael’s Hospital, Unity Health Toronto 209 Victoria Street, 3rd Floor, Toronto, Ontario, Canada, M5B 1T8; **Alternate Corresponding Author:** Dr. Sharmistha Mishra, Rm 315, 3rd Floor, St. Michael’s Hospital, Unity Health Toronto 209 Victoria Street, 3rd Floor, Toronto, Ontario, Canada, M5B 1T8.

**Keywords:** vaccine effectiveness, cohort study, test-negative design, collider bias, selection bias, residual confounding

## Abstract

Observational studies are essential for measuring vaccine effectiveness. Recent research has raised concerns about how a relationship between testing and vaccination may affect estimates of vaccine effectiveness against symptomatic infection (symptomatic VE). Using an agent-based network model and SARS-CoV-2 as an example, we investigated how differences in the likelihood of testing by vaccination could influence estimates of symptomatic VE across two common study designs: retrospective cohort and test-negative design. First, we measured the influence of unequal testing on symptomatic VE estimates across study designs and sampling periods. Next, we investigated if the magnitude of bias in VE estimates from unequal testing was shaped by the level of immune escape (vaccine efficacy against susceptibility and against infectiousness) and underlying epidemic potential (probability of transmission). We found that unequal testing led to larger biases in the cohort design than the test-negative design and that biases were largest with lower efficacy against susceptibility. We also found the magnitude of bias was moderated by the sampling period, efficacy against infectiousness, and probability of transmission, with more pronounced moderating effects in the test-negative design. Our study illustrates that VE estimates across study designs require careful interpretation, especially in the presence of epidemic and immunological heterogeneity.

## Introduction

In real-world settings, observational studies are used to infer causal effects of how well vaccines protect against infection^1,2^. Accurate estimates of vaccine effectiveness (VE) are important as they are used to evaluate the impact of vaccines at a population-level and can affect vaccine confidence^3^. Most VE observational studies use population-level data collected through passive surveillance systems that rely on tests conducted to screen for, or diagnose, infections. However, testing for a given infection depends on many factors: variability in recommendations across populations and risk factors for testing and treatment; access and uptake shaped in part by healthcare engagement; underlying burden of alternate etiologies of symptoms that drive testing. When some of these factors also influence vaccination, they can create unequal rates of testing by vaccination status, introducing biases into VE estimates such as residual confounding (due to unmeasured confounders), or selection bias due (when restricting study samples to those tested)^4–7^. Concerns surrounding biases in VE estimates due to unequal testing by vaccination status were brought to the forefront during the SARS-CoV-2 pandemic, based on empirical data that revealed associations between vaccination status with testing^8,9^. Quantifying the direction and magnitude of biases in VE estimates due to unequal testing by vaccination, and the conditions under which these biases are most influential, could inform interpretation of VE estimates and study design to reduce biases.

Two observational study designs commonly used to estimate VE are the retrospective cohort (e.g.,^10–14)^ and retrospective test-negative designs (e.g.,^15–19)^. Cohorts use vaccination status to assign exposure and estimate the relative risks of infection (between the vaccinated and unvaccinated cohorts) that may occur over a given sampling period. Classification of the outcome (infected vs. not infected) depends on testing for infection. Therefore, unmeasured confounders that lead to unequal testing by vaccination status can cause residual confounding in VE estimates from the cohort design^4,20,21^.

Test-negative designs restrict the study sample only to individuals who received a test for the infection in an attempt to eliminate unmeasured confounding^22,23^. At a given point in the epidemic (sampling period), cases (individuals who tested positive) and controls (individuals who tested negative) are compared to determine if the distribution of the exposure (vaccination) differs between cases and controls, using odds ratios^2^. However, as described by Westreich and Hudgens^6^ and Sullivan, Tchetgen Tchetgen, and Cowling^7^ through a directed acyclic graph (DAG), restricting the study sample to tested individuals may introduce a selection bias via selecting on a collider (also referred to as collider bias). As the exposure (vaccination) and outcome (infection) both affect the likelihood of being tested, selecting on testing distorts the association between vaccination and infection, thereby creating biased VE estimates^6,7^.

In an attempt to eliminate the selection bias due to selecting on testing, some test-negative design studies use symptomatic infection as the outcome to estimate the vaccine’s protection against symptomatic infection (hereafter referred to as symptomatic VE) (e.g.,^15,24,25^). Here, these studies also restrict their sample to only those individuals with symptoms, which attempts to block the association between infection and testing to mitigate the collider bias. However, we posit that selecting on symptoms may not fully block the association between infection and testing, therefore allowing the collider bias to persist.

Various studies have qualitatively described the mechanisms by which unequal testing by vaccination could lead to residual confounding or selection bias^6,7^. However, less is known about the magnitude of bias in VE estimates due to unequal testing by vaccination in cohort vs. test-negative designs; and importantly, the epidemic and immunological properties under which biases in VE estimates are amplified or dampened in the presence of unequal testing.

Epidemic and immunological properties have the potential to shape biases in VE over time and across epidemics. Epidemic properties such as the force of infection (incidence per susceptible) and the proportion of the population that are susceptible due to infection-acquired immunity can vary over the course of an epidemic and thus can influence the degree of bias in VE estimates over time. Epidemic and immunological properties that determine the overall size and length of an epidemic may also vary with changing contexts and evolving pathogens, potentially shaping biases across epidemics. One epidemic condition that can vary is the underlying epidemic potential (probability of transmission), which could differ depending on underlying contact rates and/or biological properties of the virus (e.g. increased transmissibility of SARS-CoV-2 variants^26,27^. Immunological properties such as the biological efficacy of vaccines against susceptibility (individual-level protection against acquiring infection^28^) and/or against infectiousness (individual-level reduction in transmitting virus if infected^28^) can also vary across different phases of an epidemic as seen with the increased immune escape of SARS-CoV-2 variants^29^. As such, epidemic and immunological properties that determine the overall size and length of an epidemic may moderate the magnitude of bias observed in a given epidemic.

In this study, we developed an agent-based network model, using SARS-CoV-2 as an example, to examine the magnitude of bias in symptomatic VE estimates due to unequal testing by vaccination status across retrospective cohort and test-negative designs. Here our causal relationship of interest is the effect of vaccination on symptomatic SARS-CoV-2 infection (Fig. 1). First, using our simulated data, we compared biases produced in the two study designs at the same sampling period and then across different lengths of the sampling period within the same study design (Objective 1). We then assessed the extent to which biases were moderated by immunological properties (vaccine efficacy against susceptibility and against infectiousness) (Objective 2); and by the underlying epidemic potential (probability of transmission) (Objective 3).

**Fig. 1:**
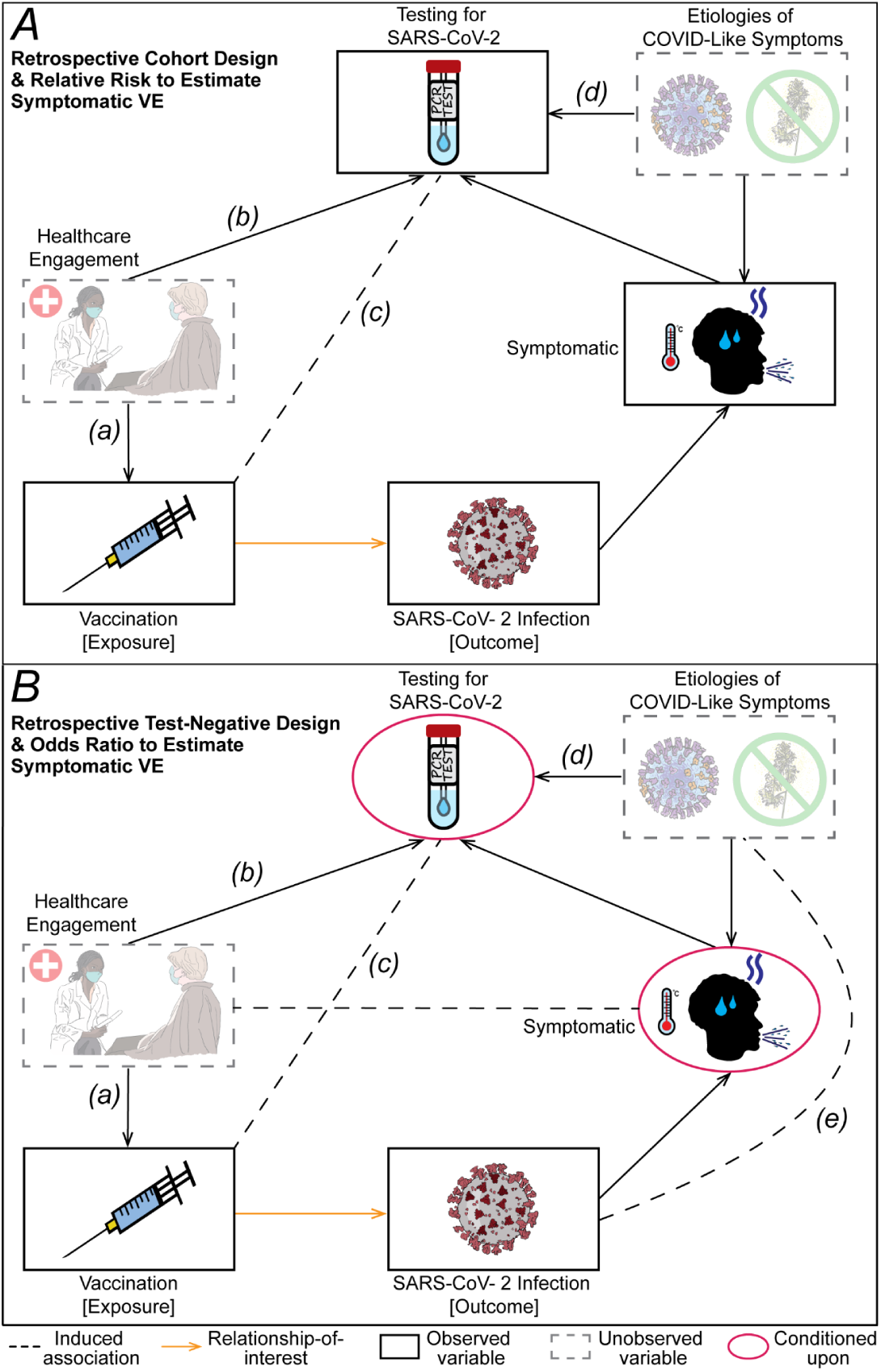
Directed Acyclic Graphs (DAGs) for estimating vaccine effectiveness against symptomatic SARS-CoV-2 infection (symptomatic VE) using a retrospective cohort design with relative risk (A) and a retrospective test-negative design with an odds ratio (B). The DAG depicts causal relationships (arrows) and induced associations (dashed lines). As the vaccine does not decrease the likelihood of developing symptoms, effectiveness against symptomatic infection is equivalent to effectiveness against infection (yellow arrow). Causal relationships between healthcare engagement with vaccination (a) and with testing for SARS-CoV-2 (b) induces an association between vaccination and testing for SARS-CoV-2 (c). As all individuals who are not symptomatic do not have etiologies of COVID-like symptoms and therefore do not get tested, an association also exists between testing for SARS-CoV-2 and etiologies of COVID-like symptoms (d) (depicted in B). Conditioning on symptomatic blocks the association between infection and testing via symptomatic; however it also induces an association between SARS-COV-2 infection and etiologies of COVID-like symptoms (e) through which infection is associated with testing.

## Results

### Comparison of biases from testing differences across study designs and sampling periods

Using an agent-based network model and our default parameters (Fig. 2; Table 1), we simulated three testing scenarios that varied the magnitude of the relationship between testing and vaccination (“c” in Fig. 1): equal testing scenario (equal probability of testing by vaccination status); moderately unequal testing scenario (vaccinated with 1.76 times higher testing); and highly unequal testing scenario (vaccinated with 2.36 times higher testing). We assumed testing could only occur if individuals had symptoms from SARS-CoV-2 infection or had COVID-like symptoms from alternate etiologies. We then measured the magnitude of bias in VE estimates produced by the cohort design (*VE*_*RR*_) and the test-negative design (*VE*_*OR*_) and compared them across study designs and sampling periods. We ran the three testing scenarios using both a higher and lower efficacy against susceptibility (0.55 and 0.1), which was equivalent to higher and lower true symptomatic VE (see Methods for details).

**Fig. 2:**
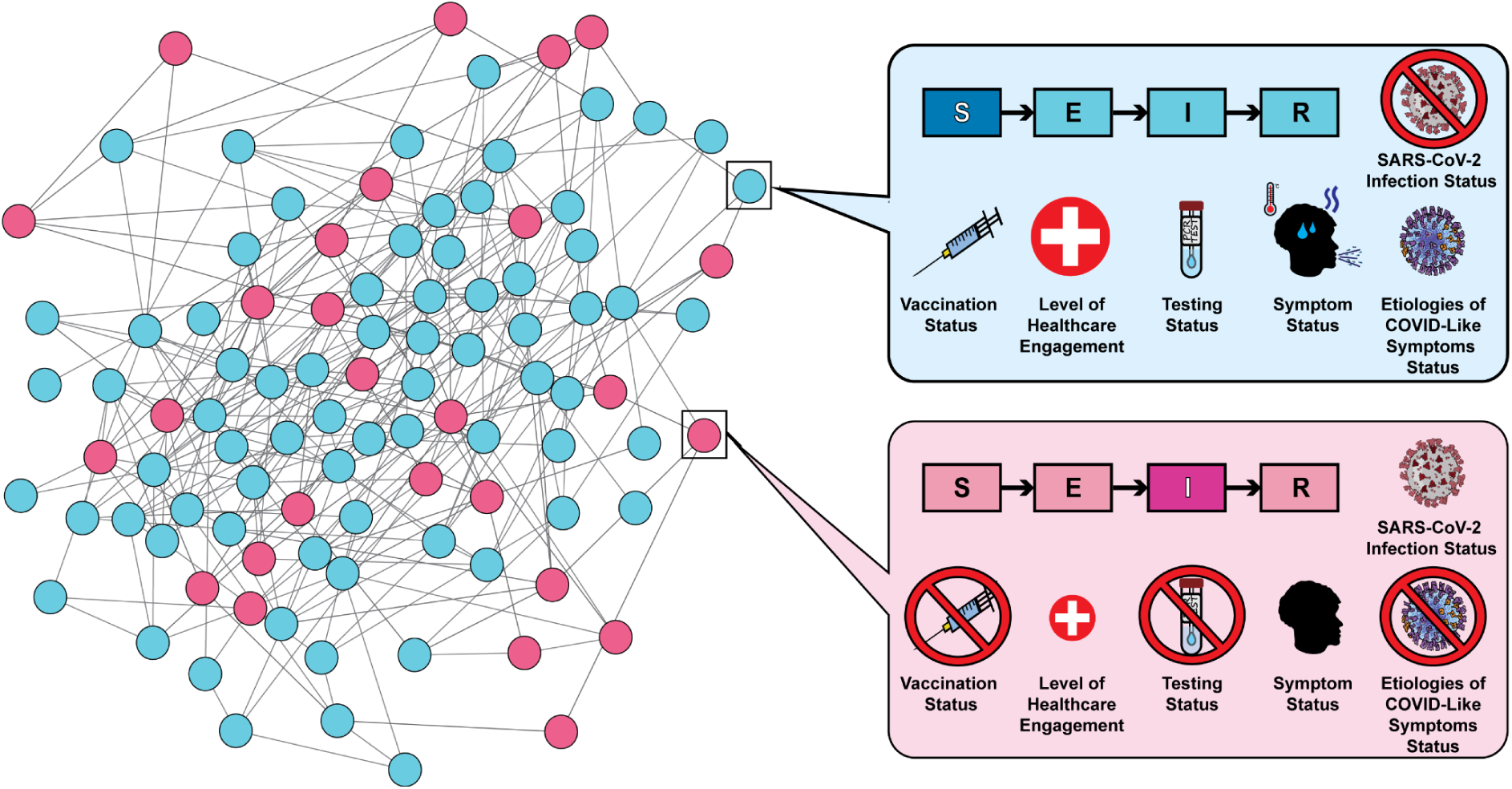
Example of the agent-based network model used to generate SARS-CoV-2 transmission dynamics. Nodes represent individuals who are vaccinated (blue) and unvaccinated (pink); edges represent contact that allows for respiratory transmission. Each node also has individual-level attributes including SARS-CoV-2 infection status (Susceptible [never infected], Exposed, Infectious; Recovered [previously infected]), vaccination status (vaccinated; unvaccinated), level of healthcare engagement (low; high), testing status (untested; tested positive for SARS-CoV-2; tested negative for SARS-CoV-2), symptom status (symptomatic; asymptomatic/no symptoms), and etiologies of COVID-like symptoms status (exposed to alternate etiologies of COVID-like symptoms; unexposed to alternate etiologies of COVID-like symptoms).

**Table 1:**
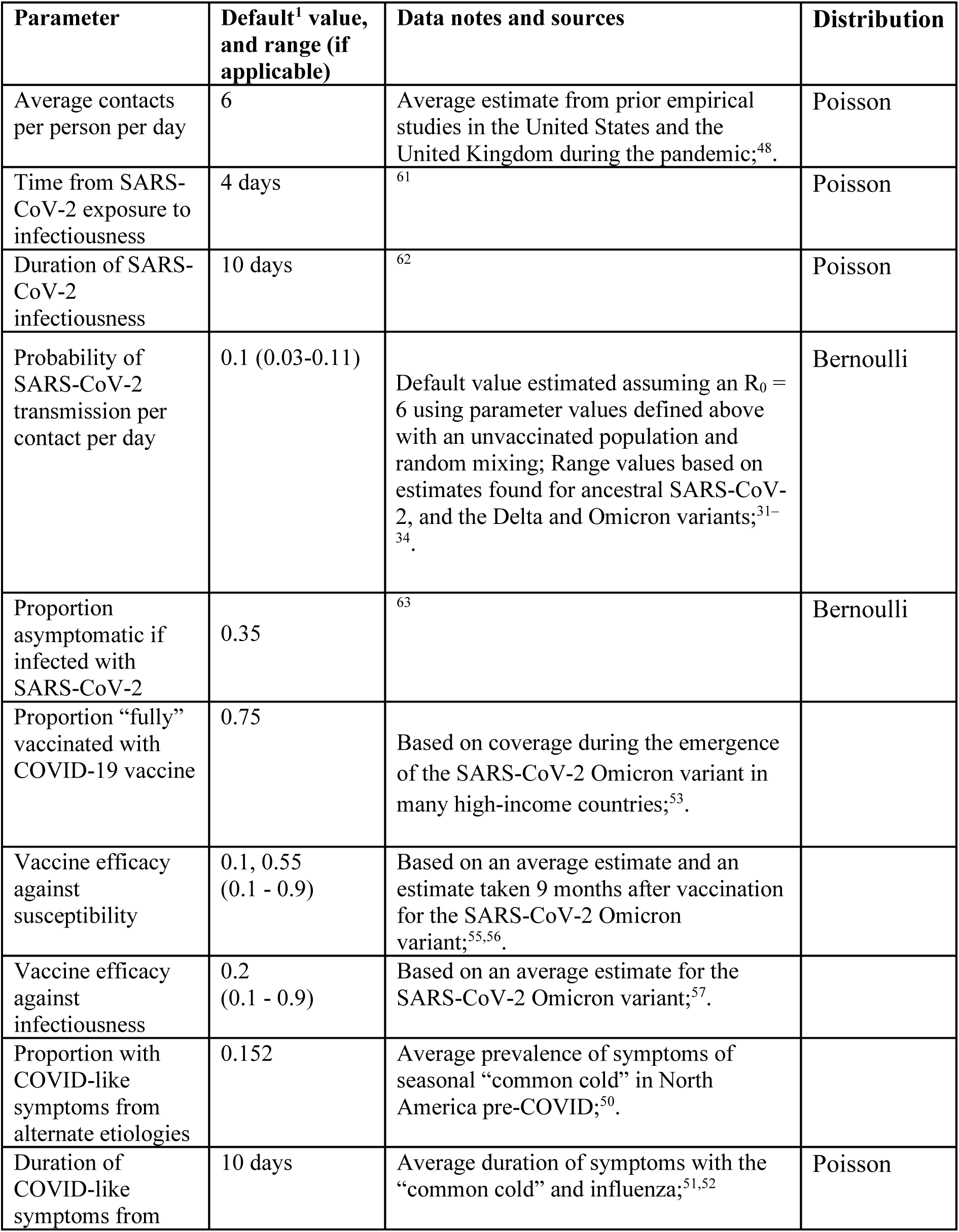
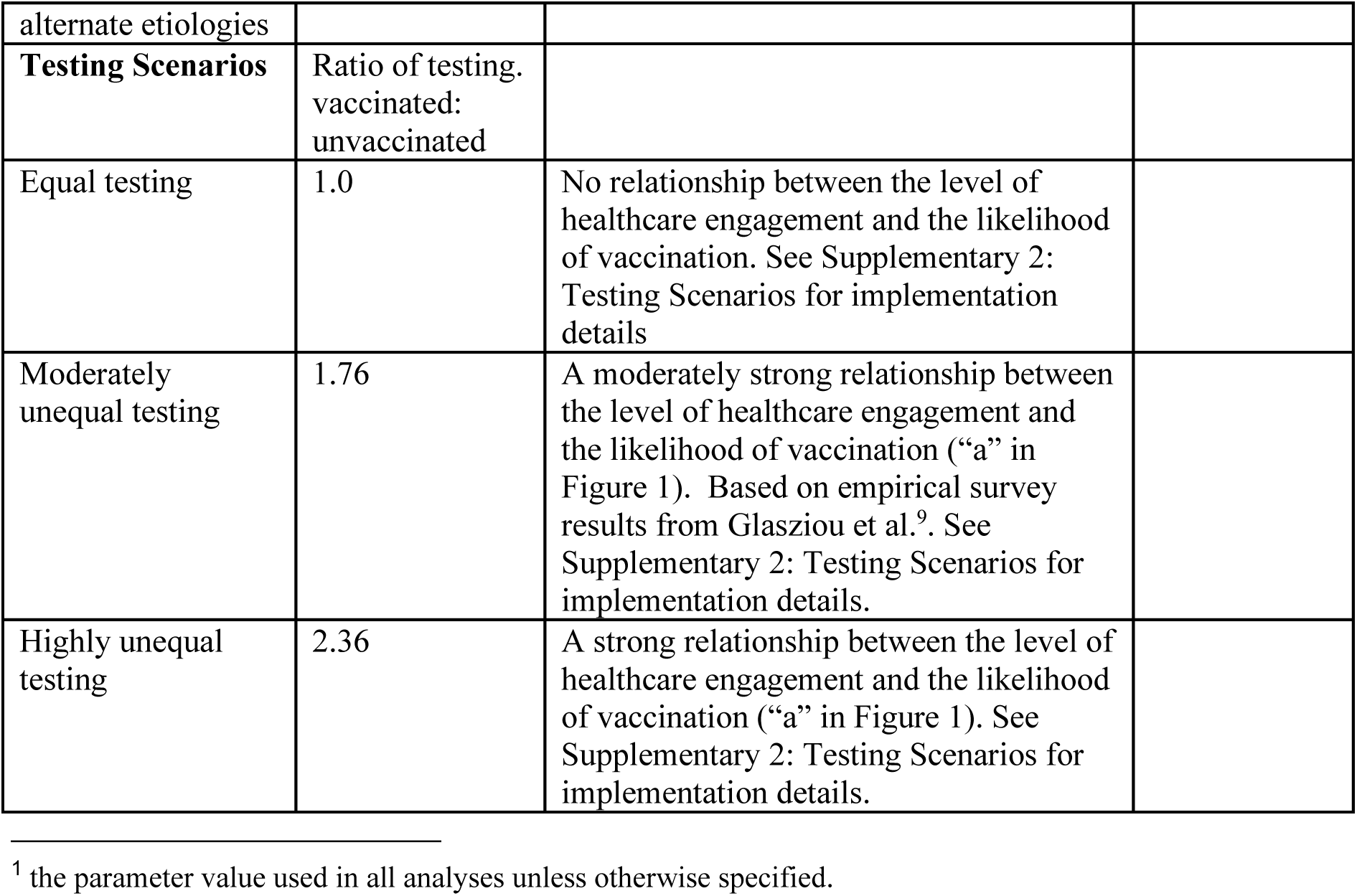
Parameter values for agent-based model including default values, reference(s) for the parameter values and the range of sensitivity vales for those parameters of interest.

| Parameter | Default<sup>1</sup> value, and range (if applicable) | Data notes and sources | Distribution |
| --- | --- | --- | --- |
| Average contacts per person per day | 6 | Average estimate from prior empirical studies in the United States and the United Kingdom during the pandemic;<sup>48</sup>. | Poisson |
| Time from SARS-CoV-2 exposure to infectiousness | 4 days | <sup>61</sup> | Poisson |
| Duration of SARS-CoV-2 infectiousness | 10 days | <sup>62</sup> | Poisson |
| Probability of SARS-CoV-2 transmission per contact per day | 0.1 (0.03-0.11) | Default value estimated assuming an $R_0 = 6$ using parameter values defined above with an unvaccinated population and random mixing; Range values based on estimates found for ancestral SARS-CoV-2, and the Delta and Omicron variants;<sup>31–34</sup>. | Bernoulli |
| Proportion asymptomatic if infected with SARS-CoV-2 | 0.35 | <sup>63</sup> | Bernoulli |
| Proportion “fully” vaccinated with COVID-19 vaccine | 0.75 | Based on coverage during the emergence of the SARS-CoV-2 Omicron variant in many high-income countries;<sup>53</sup>. |  |
| Vaccine efficacy against susceptibility | 0.1, 0.55 (0.1 - 0.9) | Based on an average estimate and an estimate taken 9 months after vaccination for the SARS-CoV-2 Omicron variant;<sup>55,56</sup>. |  |
| Vaccine efficacy against infectiousness | 0.2 (0.1 - 0.9) | Based on an average estimate for the SARS-CoV-2 Omicron variant;<sup>57</sup>. |  |
| Proportion with COVID-like symptoms from alternate etiologies | 0.152 | Average prevalence of symptoms of seasonal “common cold” in North America pre-COVID;<sup>50</sup>. |  |
| Duration of COVID-like symptoms from | 10 days | Average duration of symptoms with the “common cold” and influenza;<sup>51,52</sup> | Poisson |

<sup>1</sup> the parameter value used in all analyses unless otherwise specified.

|  |  |  |
| --- | --- | --- |
| alternate etiologies |  |  |
| **Testing Scenarios** | Ratio of testing.<br>vaccinated:<br>unvaccinated |  |
| Equal testing | 1.0 | No relationship between the level of healthcare engagement and the likelihood of vaccination. See Supplementary 2: Testing Scenarios for implementation details |
| Moderately unequal testing | 1.76 | A moderately strong relationship between the level of healthcare engagement and the likelihood of vaccination (“a” in Figure 1). Based on empirical survey results from Glasziou et al.<sup>9</sup>. See Supplementary 2: Testing Scenarios for implementation details. |
| Highly unequal testing | 2.36 | A strong relationship between the level of healthcare engagement and the likelihood of vaccination (“a” in Figure 1). See Supplementary 2: Testing Scenarios for implementation details. |

Under equal testing, the cohort design produced unbiased *VE*_*RR*_ estimates that were stable over the successive sampling periods (Fig. 3a,b). Under equal testing, the test-negative design produced unbiased *VE*_*OR*_ estimates in early sampling periods (e.g. day 20) at the beginning of the epidemic (similar to *VE*_*RR*_) (Fig. 3e,f). As the epidemic progressed, there were a larger number of cumulative symptomatic infections (Fig. 3c,d). Consequently, the test-negative design overestimated true symptomatic VE, aligning with the established knowledge that the odds ratio (used to calculate *VE*_*OR*_) underestimates the relative risk (used to calculate *VE*_*RR*_ as the outcome becomes more frequent^30^.

**Fig. 3:**
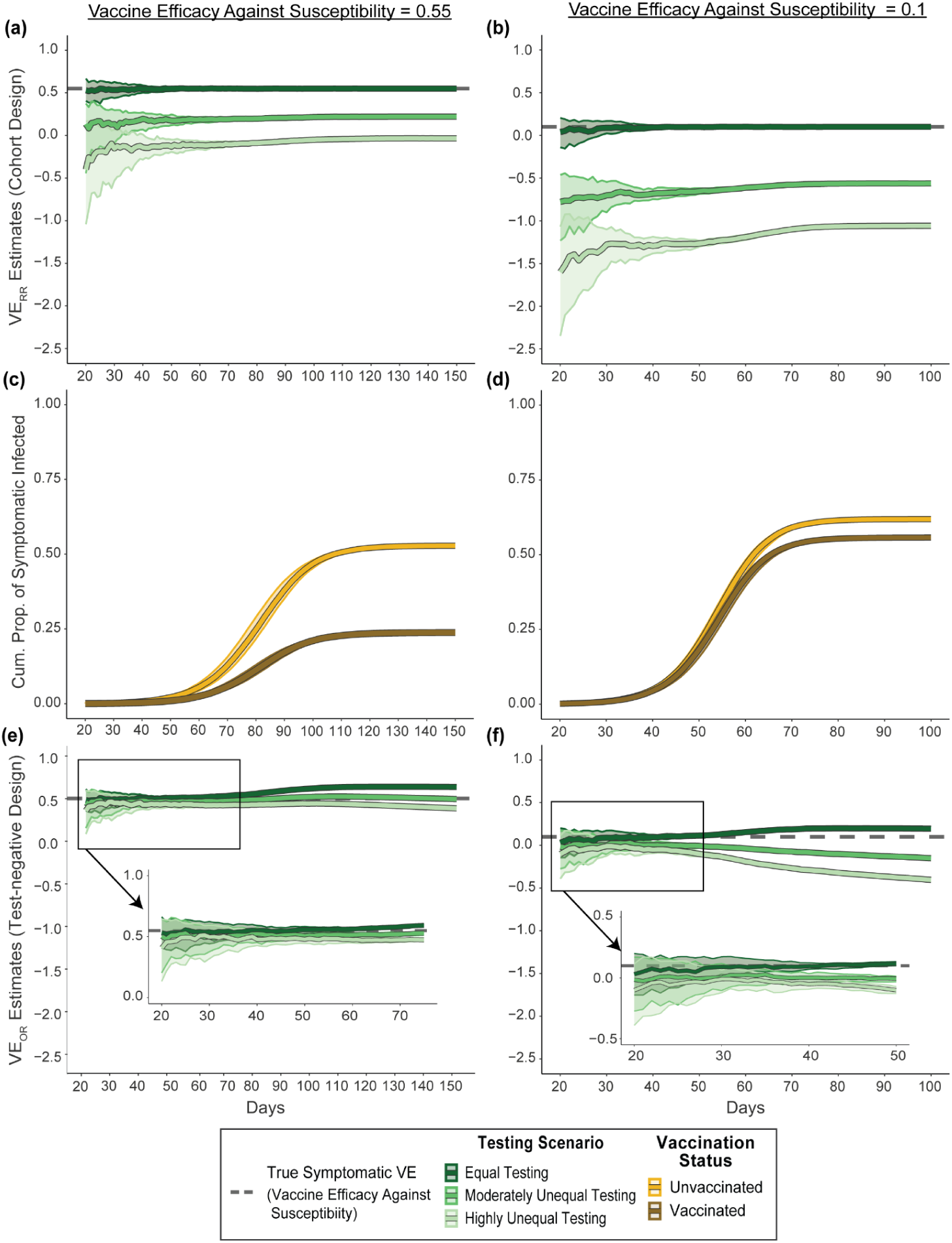
Symptomatic vaccine effectiveness estimates across a range of sampling periods from the retrospective cohort design (*VE*_*RR*_, a and b) and the retrospective test-negative design (*VE*_*OR*_, e and f); and the cumulative proportion of symptomatic infections by vaccination status (c and d). The true symptomatic VE (i.e. the level of vaccine efficacy against susceptibility) is depicted by the grey dashed line. Symptomatic VE estimates (*VE*_*RR*_ and *VE*_*OR*_) were calculated across three testing scenarios (green): equal testing by vaccination status, moderately unequal testing (vaccinated with 1.76 times higher testing), and highly unequal testing (vaccinated with 2.36x higher testing). Cumulative proportion of symptomatic infection represents the cumulative level of symptomatic infection for unvaccinated (yellow) and vaccinated (dark gold). Central lines depict the median across 100 epidemic realizations and the shaded area represents the interquartile range.

Under unequal testing, the cohort design produced *VE*_*RR*_ that underestimated the true symptomatic VE (Fig. 3a,b). Over the course of the epidemic (and thus longer sampling periods), the magnitude of the underestimate decreased and then stabilised (Fig. 3a,b). This decrease coincided with unvaccinated individuals being tested later than vaccinated individuals (Supplementary 1: Timing of Testing). Stabilization of the pattern occurred at the end of the epidemic when there were no more individuals with SARS-CoV-2 available for testing (Fig. 3c,d). The magnitude of the underestimates was larger in the highly unequal testing scenario versus the moderately unequal testing scenario (Fig. 3a,b).

Under unequal testing, the test-negative design also produced *VE*_*OR*_ that underestimated the true symptomatic VE with larger biases occurring in the highly unequal testing scenario (Fig. 3e,f). However, the test-negative design produced smaller biases than the cohort design with the difference in the magnitude of bias between study designs most pronounced when true symptomatic VE (i.e. efficacy against susceptibility) was low.

In contrast to the cohort design, biases grew larger over the sampling period in the test-negative design. This pattern arose with the test-negative design due to greater differential outcome misclassification by vaccination status that occurred with successive sampling periods (Supplementary Fig. 1).The opportunities for misclassification grew because there were more opportunities for individuals to develop COVID-like symptoms from alternate etiologies at later sampling periods and to be erroneously classified (e.g. more individuals with both prior undiagnosed SARS-CoV-2 infections and COVID-like symptoms are available to test “negative” and be classified as controls; Supplementary 1: Misclassification Example; Supplementary Fig. 2).

Irrespective of the sampling period, the magnitude of bias from the test-negative design was amplified in the moderately and highly unequal testing scenarios when there was a higher prevalence of COVID-like symptoms from alternate etiologies (Supplementary Fig. 3). Higher prevalence of COVID-like symptoms from other etiologies increased the potential for individuals to receive a negative test, allowing for more potential misclassifications to occur. Varying the prevalence of COVID-like symptoms from alternate etiologies had no influence on the magnitude of bias in estimates from the cohort design.

### Influence of immunological properties on biases from testing differences

To determine whether varying immunological properties could shape the magnitude of biases from testing differences, we repeated the above analyses but now varied the levels of efficacy against susceptibility (0.1 to 0.9) and efficacy against infectiousness (0.1 to 0.9). Here, we opted to use a single time point (the highest SARS-CoV-2 epidemic growth point [inflection point]) so that VE estimates could be easily compared across levels of vaccine efficacies.

We found that efficacy against susceptibility had a large influence on the magnitude of bias in both study designs (Fig. 4). Both designs were also capable of producing negative estimates of the true symptomatic VE when efficacy was low, even with moderately unequal testing (Fig. 3e,f). The moderating effect of efficacy against susceptibility was more pronounced as testing by vaccination changed from moderately to highly unequal.

**Fig. 4:**
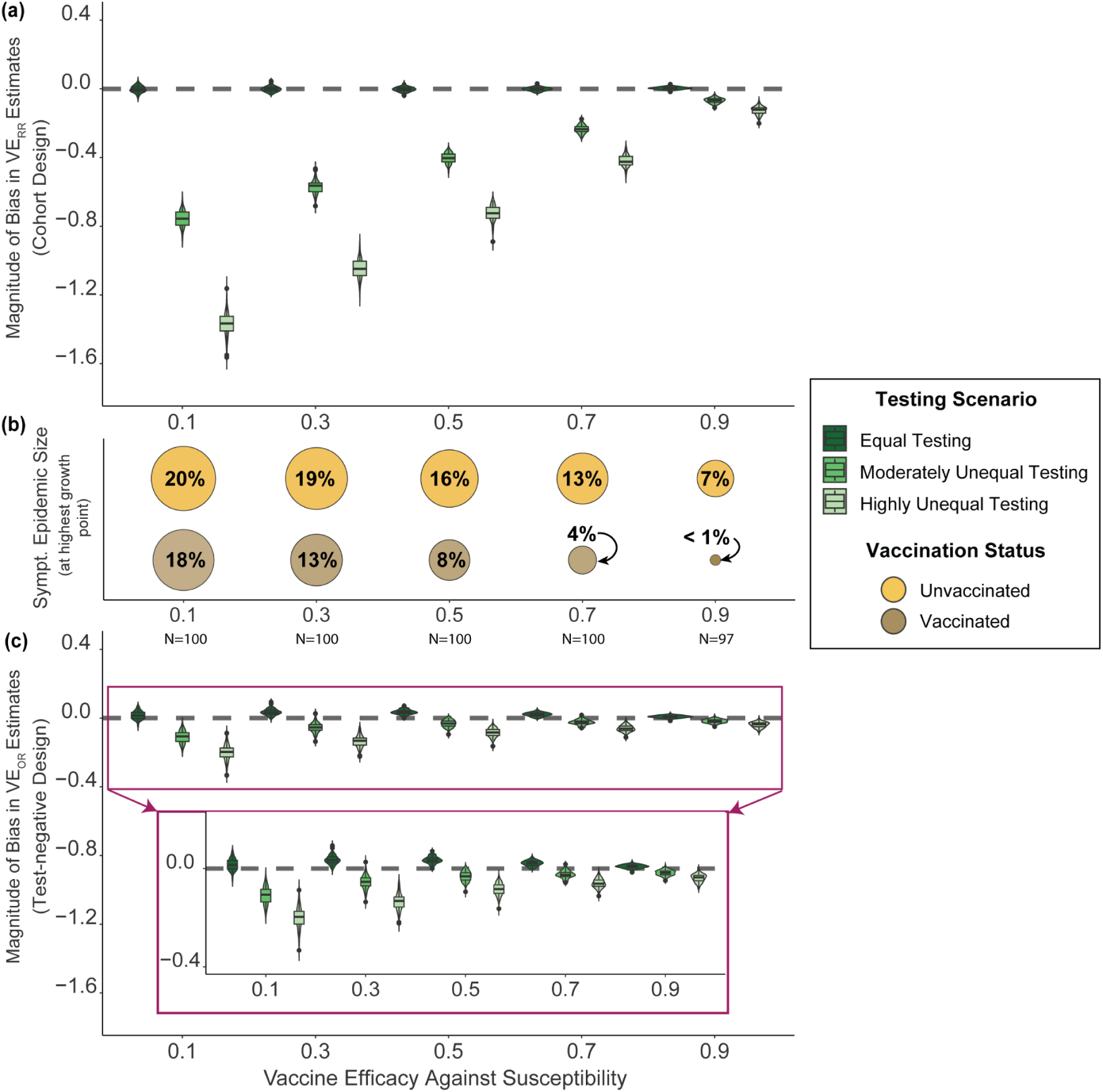
Magnitude of bias in symptomatic vaccine effectiveness (symptomatic VE) estimates and the epidemic size by testing scenarios and levels of vaccine efficacy against susceptibility. The magnitude of bias (how much estimates underestimated the vaccine’s “true” protection against symptomatic infection) was calculated across three testing scenarios (green) - equal testing by vaccination status, moderately unequal testing (vaccinated with 1.76 times higher testing), and highly unequal testing (vaccinated with 2.36x higher testing) - for the retrospective cohort design (*VE*_*RR*_) (a) and the retrospective test-negative design (*VE_OR_*) (c). The epidemic size - the cumulative proportions of individuals with symptomatic SARS-CoV-2 who were unvaccinated (yellow) and vaccinated (dark gold) (b) - and VE estimates were each sampled at the highest positive epidemic growth point per epidemic (the inflection point). Each epidemic scenario was simulated 100 times; simulated epidemics with less than 0.1% cumulative infections were removed (N= the # of epidemic realizations per scenario).The median time of sampling by the level of vaccine efficacy against susceptibility was *t =* 50 for 0.1, *t =* 58 for 0.3, *t =* 68 for 0.5, *t =* 92 for 0.7, and *t =* 185 for 0.9.

Efficacy against infectiousness also had a moderating effect on the magnitude of underestimates caused by unequal testing in both study designs (Fig. 5). The moderating effect was less pronounced than that of efficacy against susceptibility. The moderating effect of efficacy against infectiousness was present across both high and low efficacy against susceptibility.

**Fig. 5:**
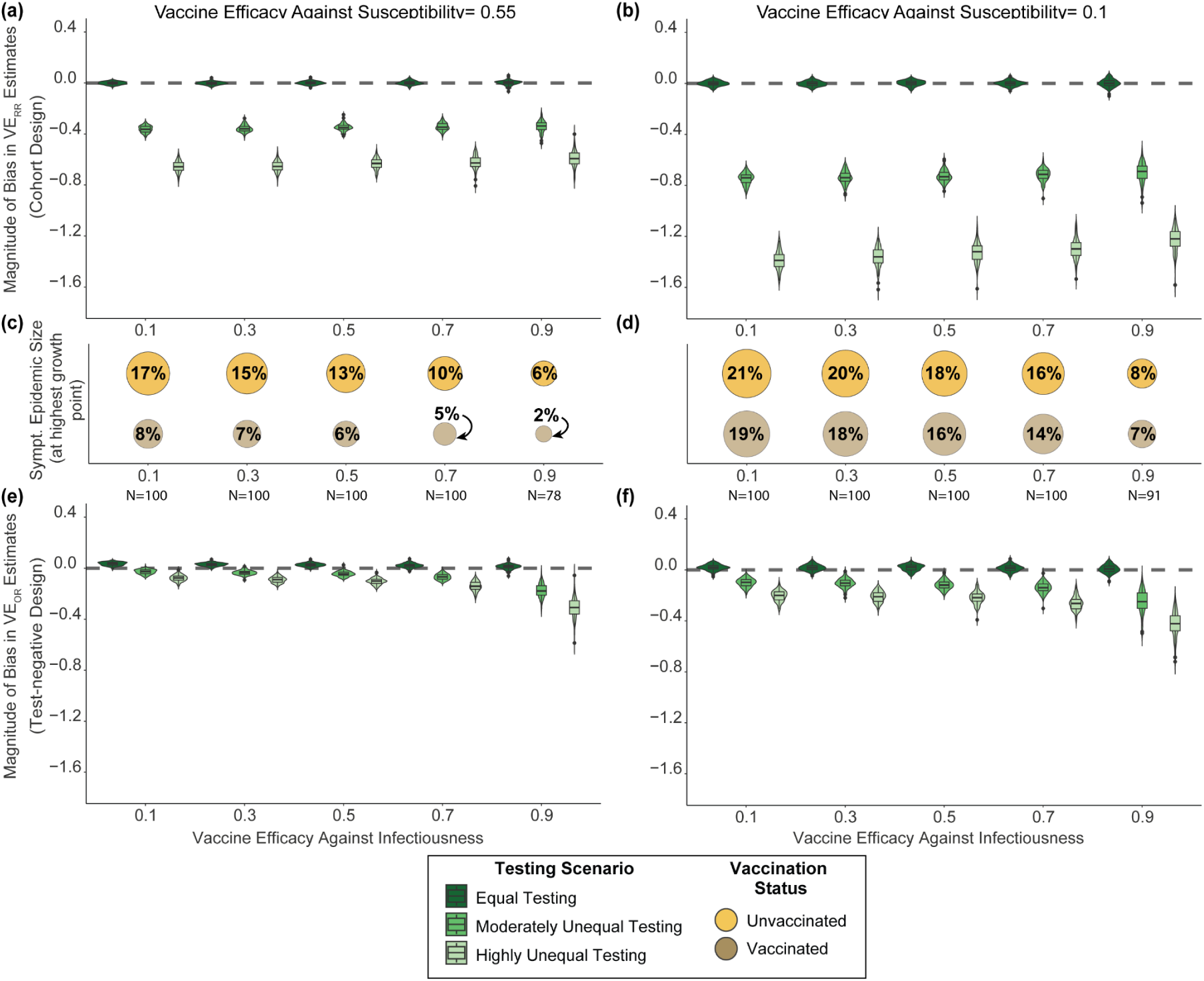
Magnitude of bias in symptomatic vaccine effectiveness (symptomatic VE) estimates and the epidemic size by testing scenarios and levels of vaccine efficacy against infectiousness given higher and lower vaccine efficacy against susceptibility (0.55 and 0.1). The magnitude of bias (how much estimates underestimated the vaccine’s “true” protection against symptomatic infection) was calculated across three testing scenarios (green) - equal testing by vaccination status, moderately unequal testing (vaccinated with 1.76 times higher testing), and highly unequal testing (vaccinated with 2.36x higher testing) - for the retrospective cohort design (*VE*_*RR*_) (a and b) and the retrospective test-negative design (*VE*_*OR*_) (e and f). The epidemic size - the cumulative proportions of individuals with symptomatic SARS-CoV-2 who were unvaccinated (yellow) and vaccinated (dark gold) (c and d) - and symptomatic VE estimates were each calculated at the point of the highest positive epidemic growth per epidemic (the inflection point). Each epidemic scenario was simulated 100 times; simulated epidemics with less than 0.1% cumulative infections were removed (N= the # of epidemic realizations per scenario). When vaccine efficacy against susceptibility was higher (0.55), the median time of sampling across vaccine efficacy against infectiousness was *t =* 69 for 0.1, *t =* 79 for 0.3, *t =* 93 for 0.5, *t =* 123 for 0.7, and *t =* 290 for 0.9; when vaccine efficacy against susceptibility was lower (0.1), the median time of sampling was *t =* 48 for 0.1, *t =* 54 for 0.3, *t =* 64 for 0.5, *t =* 85 for 0.7, and *t =* 167 for 0.9.

Under unequal testing, the moderating effect of each mechanism of efficacy (against susceptibility, against infectiousness) also varied by study design. With differences across efficacy against susceptibility, the moderating effect was more pronounced with the cohort design than the test-negative design (noting also that the absolute magnitude of the bias was much smaller with the test-negative design) (Fig. 4a,c). With differences in efficacy against infectiousness, the moderating effect was more pronounced with the test-negative design than the cohort design and was most evident under scenarios of high efficacy against infectiousness (Fig. 5a,b,e,f).

Efficacy against susceptibility had a larger influence than efficacy against infectiousness, and a different influence on the magnitude of bias by study design, because of the differential effect that these efficacies had on the cumulative numbers of symptomatic infections by vaccination status - the main components of VE calculations. Although higher efficacy against susceptibility and higher efficacy against infectiousness resulted in longer epidemic periods with smaller epidemic sizes (Supplementary Fig. 4-6), changes in efficacy against susceptibility led to differential reductions in the cumulative proportions of symptomatic infections by vaccination status (Fig. 4b; Supplementary Fig. 4) while changes in efficacy against infectiousness led to more proportionate reductions by vaccination status (Fig. 5c,d; Supplementary Fig. 5-6). Specifically, increasing efficacy against susceptibility caused symptomatic infections among vaccinated to shrink more than among the unvaccinated, such that the absolute difference between these two groups (vaccinated versus unvaccinated) tended to monotonically increase until reaching very high levels of efficacy (Fig. 4b; Supplementary Fig. 4). The absolute difference was amplified when testing was more unequal. In contrast, changes in efficacy against infectiousness caused a similar reduction in the cumulative proportions of symptomatic infections by vaccination status as vaccinated and unvaccinated individuals both benefited from the reduction of infectiousness provided by those who were vaccinated (Fig. 5c,d; Supplementary Fig. 5-6). The consequence of this on the calculations of relative risk and odds ratios explain the differences in the moderating effects by study design and are detailed in the supplementary information with examples (Supplementary 1: Moderating Effect Calculations; Supplementary Fig. 7-10).

### Influence of underlying epidemic potential (probability of transmission) on biases from testing differences

To determine whether changing epidemic properties could shape the magnitude of biases due to testing scenarios, we varied the underlying epidemic potential - the probability of transmission (0.033 to 0.11^31–34)^ and assessed the effect on biases in VE estimates. We again used a single time point (the highest SARS-CoV-2 epidemic growth point [inflection point]).

Similar to the effect of efficacy against infectiousness, decreasing the probability of transmission led to a similar, proportionate, reduction in the cumulative proportions of symptomatic infections by vaccination status (Fig. 6c,d; Supplementary Fig. 11-12). As such, the moderating influence of the probability of transmission demonstrated a similar pattern to that of efficacy against infectiousness. In the context of high transmission, changes in the transmission had a negligible influence on the magnitude of bias produced in both study designs (similar to the pattern observed when efficacy against infectiousness was low) (Fig. 6a,b,e,f). However, in the context of low transmission, small changes in transmission led to larger changes in the magnitude of bias but only in the test-negative design (similar to the pattern observed when efficacy against infectiousness was high) (Fig. 6e,f).

**Fig. 6:**
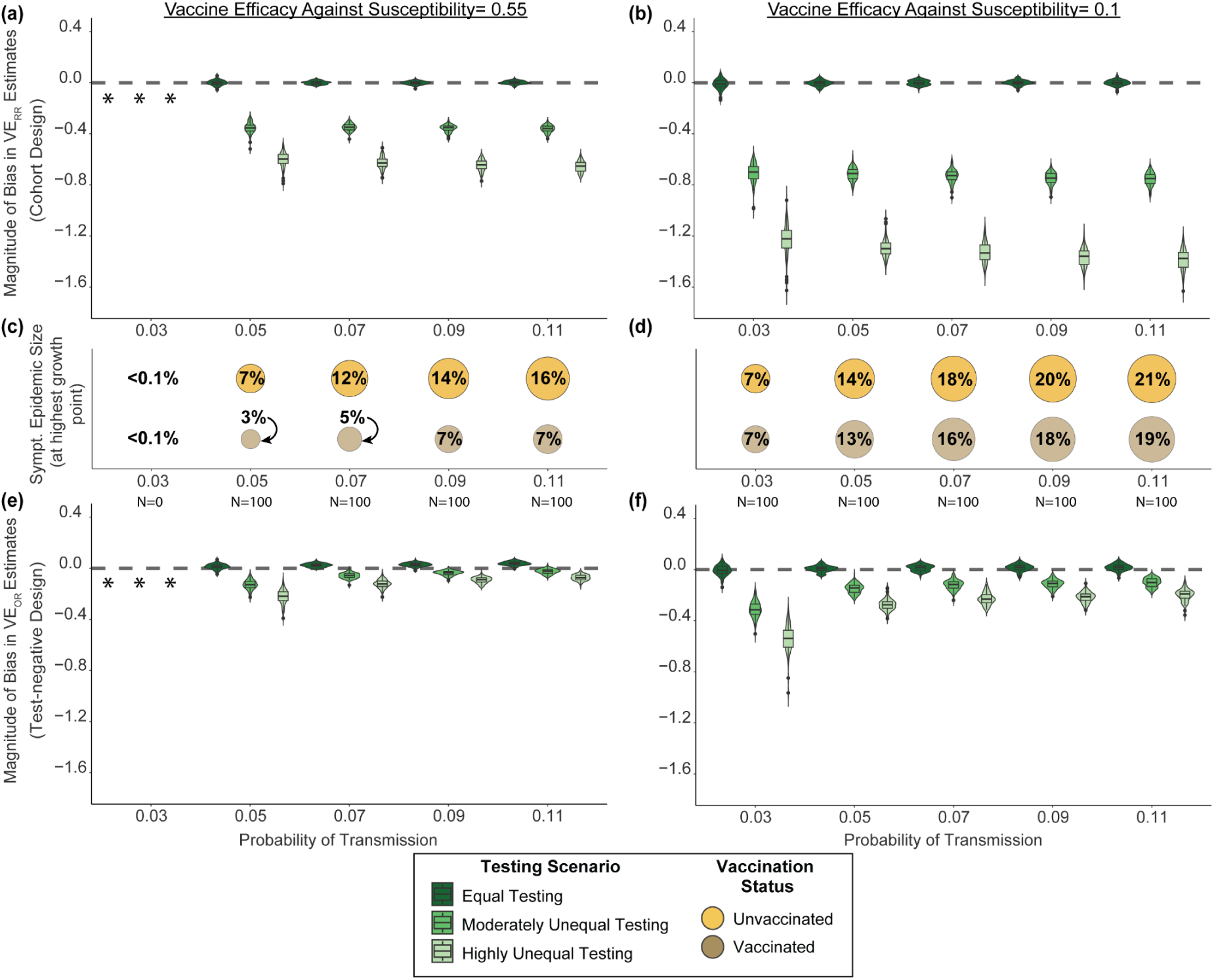
Magnitude of bias in symptomatic vaccine effectiveness (symptomatic VE) estimates and the epidemic size by testing scenarios and probability of transmission for higher and lower vaccine efficacy against susceptibility (0.55 and 0.1). The magnitude of bias (how much estimates underestimated the vaccine’s “true” protection against symptomatic infection) was calculated for three testing scenarios (green) - equal testing by vaccination status, moderately unequal testing (vaccinated with 1.76 times higher testing), and highly unequal testing (vaccinated with 2.36x higher testing) - for the retrospective cohort design (*VE*_*RR*_) (a and b) and the retrospective test-negative design (*VE*_*OR*_) (e and f). The epidemic size - the cumulative proportions of individuals with symptomatic SARS-CoV-2 who were unvaccinated (yellow) and vaccinated (dark gold) (c and d) - and symptomatic VE estimates were each calculated at the point of the highest positive epidemic growth per epidemic. Each epidemic scenario was simulated 100 times; simulated epidemics with less than 0.1% cumulative infections were removed (N= the # of epidemic realizations per scenario). If the magnitude of bias in VE estimates could not be calculated, they were replaced with the “*” symbol. When vaccine efficacy against susceptibility was higher (0.55), the median time of sampling across probabilities of transmissibility was *t =* 202 for 0.05, *t =* 108 for 0.07, *t =* 82 for 0.09 and *t =*67 for 0.11; when vaccine efficacy against susceptibility was lower (0.1), the median time of sampling was *t =* 225 for 0.03, *t =* 90 for 0.05, *t =* 65 for 0.07, *t =* 54 for 0.09, and *t =* 47 for 0.11.

The difference in the direction and magnitude of biases by study design occurred because of the extent to which the ratio in the numerator (in the relative risk) changed with changes in the probability of transmission, and the extent to which the ratio in the numerator and in the denominator changed (with the odds ratio). These patterns again are similar to those observed with efficacy against infectiousness. Detailed explanation and examples of how these calculations change across probabilities of transmission and by study design are provided in the Supplementary Information (Supplementary 1: Moderating Effect Calculations; Supplementary Fig. 13-14).

## Discussion

Using simulation modelling and SARS-CoV-2 as the motivating example, we examined the magnitude of bias in symptomatic VE due to unequal testing by vaccination status and explored how epidemic and immunological properties could shape biases across two commonly adopted observational VE study designs. Overall, in the presence of unequal testing by vaccination, we found that both the cohort and test-negative design could underestimate true symptomatic VE with greater testing differences leading to larger underestimates. However, we also found that the cohort design produced larger underestimates (larger biases) than the test-negative design. The largest bias for both study designs occurred with low efficacy against susceptibility, but the moderating effect of efficacy against susceptibility on bias was more pronounced in the cohort design compared with the test-negative design. In contrast, biases produced by the test-negative design were more likely than biases produced by the cohort design to be influenced by the length of the sampling period, the efficacy against infectiousness, and the probability of transmission.

Our findings confirm and build on previous insights on the mechanisms by which unequal testing and healthcare engagement can create biased estimates of VE in cohort designs^4,20,21^ and in test-negative designs^6,7^. First, our work expands on previous qualitative studies that showcased the potential for residual confounding and selection bias with VE against infection^6,7^. We illustrated that selection bias in test-negative designs can still play an important role in affecting estimates of true symptomatic VE - although the magnitude of the bias is likely to be small. Second, our work provides a detailed comparison of the magnitude of bias found across the two most commonly-adopted study designs - the retrospective cohort and test-negative design. Our findings add support to the use of a test-negative design over that of a cohort-design in estimating VE in the presence of unequal testing by vaccination status^22^. We further identified important epidemic and immunological properties that should be considered when interpreting VE estimates from both study designs: efficacy against susceptibility, efficacy against infectiousness, and the probability of transmission.

Efficacy against susceptibility had the largest influence on the magnitude of bias in VE estimates with a more pronounced effect on estimates from the cohort design. Other studies have also found that efficacy against susceptibility can moderate the magnitude of bias produced by other mechanisms of bias such as heterogeneity in contact levels by vaccination status^35^ and confounding bias due to correlated case and control vaccination behaviours^2^. The key implication from these findings is that as a virus undergoes genetic adaptations leading to immune escape, and/or waning against susceptibility occurs, the greater the risks of larger biases being produced in VE estimates. This is likely why several studies found estimates of negative VE (suggesting that the vaccine is increasing symptomatic infection) during the emergence of the Omicron variant of SARS-CoV-2 (e.g.,^24,25^) - a variant known to have higher immune escape^36^.

There are three important implications stemming from the influence of sampling period, efficacy against infectiousness, and the epidemic potential (probability of transmission) – all of which were more pronounced with the test-negative design than with the cohort design. First, in the context of an emerging and evolving outbreak, VE studies are routinely conducted over successive time-periods. Thus, it becomes increasingly important, in the context of unequal testing, to repeat VE studies and consider the timing of sampling for interpretation of VE estimates. Early in the epidemic, a test-negative design offers a large advantage in terms of minimizing the magnitude of bias but calls for greater caution in interpretation as the epidemic evolves. Caution is particularly important given that irrespective of the sampling period, prevalence of COVID-like symptoms from alternate etiologies influenced the magnitude of bias in symptomatic VE estimates from the test-negative design. As such, the population (e.g. age-group, or prevalence of comorbidities), geography, and seasonality that govern the level of alternate etiologies for COVID-like symptoms are expected to create differences in biases with test-negative designs.

Second, although changes in efficacy against infectiousness play an overall smaller role in moderating the magnitude of biases from unequal testing compared to efficacy against susceptibility, its effects can still be prominent when using the test-negative design. Efficacy against infectiousness is therefore important to consider in the context of the test-negative design as the efficacy against infectiousness can change as vaccines are updated and as viruses, such as influenza, develop immune escape that alters their ability to transmit^37^.

Third, the finding that small changes in epidemic potential – in the context of already low epidemic potential - could have a large moderating effect on the size of biases in test-negative designs provides a key insight when conducting and comparing VE estimates across populations and geographies. This is because study settings are inherently shaped by the underlying contact network (e.g. study setting with high-density housing and occupations with high rates of contacts) which in turn, shape epidemic potential.

Taken together, when pooling or synthesizing VE estimates, future work is needed to account for sources of heterogeneity that affect the magnitude of potential biases to strengthen the robustness of evidence synthesis. In the meantime, careful inclusion of nuanced DAGs (across study designs) and cautious and transparent interpretation to communicate the potential benefits of vaccination is essential. Good practices also include anticipating and recognizing that observed VE may represent biased VE estimates in the presence of unequal testing by vaccination status. These practices are important given that addressing unequal testing remains challenging. Electronic health records in national databases typically have information on exposures and health outcomes but limited data on confounders^38^. We therefore echo the sentiments of Lewnard et al.^39^ cautioning that the test-negative design - along with other types of study designs - should not be naively applied to testing datasets collected through administrative or public health surveillance data sources. An area of future methodological work includes using externally-collected data on testing by vaccination status from representative surveys (e.g.,^8,9^) to conduct quantitative bias analysis, including simulation-based bias analysis ^40,41^.

Our simulation study has several limitations. Chief among them is our assumption that vaccination coverage was static. Rolling out a vaccine over time could potentially reduce the magnitude of bias due to unequal testing if, for example, individuals who have high health engagement (and hence are likely to be tested) also wanted to be vaccinated but were not able to immediately receive one. This effect could also be reversed if interventions reached those with the highest healthcare engagement first before being distributed to those with lower healthcare engagement (causing more unequal testing by vaccination in the beginning of the epidemic before stabilizing). Second, we assumed the severity of symptoms due to SARS-CoV-2 did not vary by vaccination status. Vaccination for SARS-CoV-2 has been shown to reduce symptoms^42,43^. Depending on the strength of the association between vaccination and testing by symptom-severity, which has been shown to affect the accuracy of VE estimates^23^, the magnitude of bias could be reduced or remain unchanged from counterbalancing associations (e.g vaccinated individuals may have milder symptoms than unvaccinated individuals, but they may also be more likely to get tested even with mild symptoms). Third, our scenarios were conditioned on vaccinated individuals testing more than unvaccinated individuals based on several surveys related to SARS-CoV-2 testing and vaccination (e.g.,^8,9^). However, unequal testing has also occurred in the opposite direction^44^, which is an important consideration when comparing VE studies across different study settings, irrespective of pathogen. If unequal testing reflects higher testing among unvaccinated individuals, the direction of the bias would change, leading to an overestimation rather than an underestimation of symptomatic VE.

Observational studies are key to assessing how well vaccines might be working in the real-world^2^. We illustrated how unequal testing by vaccination status could bias estimates of true symptomatic VE across two commonly used study designs. Overall, using a test-negative design rather than a cohort design can reduce the magnitude of biases due to unequal testing. However, interpretation of VE estimates still requires cautious interpretation, especially given heterogeneity in epidemic and immunological properties – with the most important property being vaccine efficacy against susceptibility.

## Methods

### Mechanisms of Interest

The causal relationship we are interested in is the effect of vaccination on symptomatic SARS-CoV-2 infection (Fig. 1). For simplification, we assumed that vaccination does not reduce symptoms in vaccinated individuals who acquire SARS-CoV-2 and thus the effect of vaccination on infection is the same as its effect on symptomatic infection. If infected, an individual may develop symptoms which leads to an opportunity to engage in SARS-CoV-2 testing. Both infectious and non-infectious etiologies other than SARS-CoV-2 can lead to symptoms similar to those caused by SARS-CoV-2 (referred to as etiologies of COVID-like symptoms, Fig. 1)^45^.

We assumed that the association between vaccination and testing (pathway marked as “c” in Fig. 1, which reflects unequal testing by vaccination status) is induced by a relationship between the level of healthcare engagement and vaccination (pathway marked “a” in Fig. 1) and a relationship between the level of healthcare engagement and testing (pathway marked as “b” in Fig. 1).

In the cohort design, everyone was included in the study sample. Fig. 1A depicts how unequal testing by vaccination status can lead to residual confounding in VE estimates when using the cohort design if healthcare engagement is unmeasured. In the test-negative design, the study sample was restricted to individuals who were symptomatic and tested for SARS-CoV-2 (depicted by the two “conditioned upon” circles in Fig. 1B). Conditioning on symptomatic and testing can generate a selection bias in VE estimates due to selecting/conditioning on a collider. Testing meets the criteria as a collider because both the exposure (vaccination) and the outcome (infection) are associated with testing. While conditioning on symptomatic blocks the association between SARS-CoV-2 infection and testing (as symptomatic is a mediator), it induces an association between SARS-CoV-2 infection and etiologies of COVID-like symptoms (depicted by dashed line in Fig. 1B) through which infection is associated with testing (see Supplementary 2: Selection Bias for details).

To focus our examination on mechanisms of biases due to unequal testing by vaccination status, we assumed no other sources of unmeasured confounding.

### Model Overview

We used an agent-based model to simulate SARS-CoV-dynamics and to model the individual-level variations in vaccination and testing (Fig. 2). We chose an agent-based model as it is a flexible approach that can capture emergent phenomena arising from complex interactions in a heterogeneous population^46^. In our model, each individual (i.e. agent) was assigned attributes to capture health-states related to SARS-CoV-2: Susceptible, Exposed, Infectious, Recovered and Immune (where individuals are assumed to be fully protected from re-infection). Other individual-level attributes that vary over time include: symptom status (symptomatic or asymptomatic), SARS-CoV-2 testing (tested or not tested); SARS-CoV-2 diagnosis if tested (tested positive vs negative). Attributes that do not vary over time include: healthcare engagement (low or high); and vaccination status (vaccinated or unvaccinated).

The model updates each individual’s attributes daily. Table 1 summarises the parameters and their corresponding data sources.

### Contact Patterns

Transmission occurs in daily time-steps through contacts in a homogenous and static network generated using an Erdos-Renyi random graph model^47^. The network is based on an average of 6 daily contacts^48^. Given the network is sparse, the distribution of contacts across the population follows a Poisson distribution^49^. The original placement of edges (representing contacts) is homogeneous by vaccination status, and all nodes (individuals) are connected to the main network (Supplementary 2: Contact Patterns).

### SARS-CoV-2 Transmission

Based on available data from the literature, we assigned a default value for the biological probability of transmission per contact per day, duration of the exposure state, duration of infectiousness, and proportion asymptomatic if infected with SARS-CoV-2 (Table 1). Each of these processes is stochastic and was implemented using draws from statistical distributions (Supplementary 2: SARS-COV-2 Transmission Dynamics).

### COVID-like Symptoms

At any given point in time, a stable proportion (default value of 0.152 in our primary analysis) of the simulated population experienced COVID-like symptoms from alternate etiologies^50^. We assumed symptoms lasted on average for 10 days (reflecting average duration of symptoms from the “common cold” ^51^ and influenza^52^ (Appendix 1: COVID-like Symptoms).

### Vaccination (Coverage and Mechanisms of Protection)

We applied a fixed 75% vaccination coverage^53^ such that 75% of the simulated population was fully vaccinated prior to the start of the epidemic, with no further vaccination thereafter. This reflects a scenario wherein a variant may emerge, but new or additional vaccination has not yet been rolled out.

We simulated vaccine efficacy against susceptibility to acquisition of SARS-CoV-2 and vaccine efficacy against infectiousness^54^. Efficacy against susceptibility was implemented as “all-or-nothing” protection. That is, the vaccine induced 100% protection for a subset of vaccinated individuals (set by the value of efficacy against susceptibility) and zero protection for the remainder (1-efficacy)^54^. We assumed that vaccination did not reduce the probability of becoming symptomatic.

Most of our analyses used a default value for high and low efficacy against susceptibility (0.55, informed by an average estimate of VE against Omicron infection^55^; and a default value for low efficacy against susceptibility (0.1, reflecting VE estimates taken 9 months after vaccination^56^. For vaccine efficacy against infectiousness, we used a default value of 0.2^57^.

### VE Study Design and Estimates

To mimic the retrospective cohort design in studies using population-level health administrative data, we assumed that symptomatic individuals with SARS-CoV-2 who were not tested will be misclassified as not infected when estimating true symptomatic VE using the cohort design (formula 1).

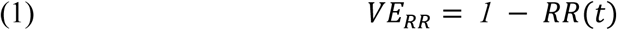

with:

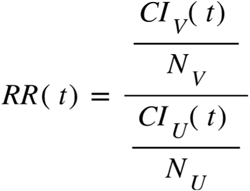

where *RR*(*t*) is a relative risk at time *t*; CI_v_(t) and CI_u_(t) are the cumulative numbers of symptomatic infected for vaccinated and unvaccinated that have been tested and diagnosed (tested positive) at time *t,* respectively; and *N*_v_ and *N*_u_ are the total numbers of vaccinated and unvaccinated individuals, respectively.

To mimic the test-negative design, we sampled from the simulated cohort individuals who were both symptomatic and tested. Using those sampled individuals we can estimate the true symptomatic VE, with formula 2:

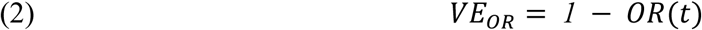

and

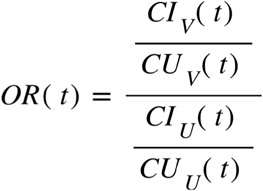

where *OR* (t) is an odds ratio at time *t*; and C_v_ (t) and C_u_ (t) are the cumulative numbers of symptomatic individuals due to etiologies of COVID-like symptoms for vaccinated and unvaccinated populations who tested negative for SARS-COV-2 and had no prior positive tests for SARS-COV-2 by time *t,* respectively.

### Testing Scenarios

We simulated three testing scenarios, each with a varying strength of the relationship between testing and vaccination (“c” Fig. 1): equal testing scenario (equal probability of testing by vaccination status); moderately unequal testing scenario (vaccinated with 1.76 times higher testing); and highly unequal testing scenario (vaccinated with 2.36 times higher testing). Here the direction of the testing differences (i.e. vaccinated individuals testing more than unvaccinated) and the magnitude of the differences in the moderately unequal testing scenario are based on empirical survey results^9^). Varying “c” also affected the timing of testing by vaccination status. Thus, in our testing scenarios, vaccinated individuals tested later than unvaccinated individuals when testing was unequal with the difference in the timing growing as unequal testing went from moderately unequal to highly unequal (Supplementary 1: Timing of Testing).

The magnitude of “c” is influenced by the likelihood of vaccination by the level of healthcare engagement (“a” in Fig. 1); and the likelihood of testing by the level of healthcare engagement (“b” in Fig. 1). We varied values of “a” to simulate the three testing scenarios above (see Supplementary 2: Testing and Healthcare Engagement and Testing Scenarios for details).

### Simulations and Analyses

Each scenario consisted of 100 epidemic realizations and each epidemic realization used a population size of 100,000. We selected these values because in scenarios using our default parameters, estimates of symptomatic VE had converged by the epidemic’s inflection point. All realizations were initialized with 10 randomly assigned individuals on day 1 of their infectiousness (See Supplementary 2: Event Scheduling). In our analyses, we estimated VE using a cohort design and test-negative design (VE_RR_ and VE_*OR*_) under the three testing scenarios. The magnitude of bias was measured as the difference between efficacy against susceptibility (the model input), and the symptomatic VE estimates produced by the cohort design (VE_RR_) and the test-negative design (VE_*OR*_). We assessed the magnitude of bias using efficacy against susceptibility as it represents the true symptomatic VE. Efficacy against susceptibility is equivalent to true symptomatic VE because 1) the vaccine provides “all-or-nothing” protection^54^; and 2) the vaccine does not reduce the likelihood of becoming symptomatic once infected. We performed model verification to confirm the accuracy of our model, and our symptomatic VE estimates (Supplementary 2: Model Verification; Supplementary Fig. 15-16). Results are reported using the median and the interquartile range from the 100 realizations.

To address how symptomatic VE can vary across study designs and across sampling periods (Objective 1), we calculated VE_RR_ and VE_*OR*_ (cumulative measurements as shown in formula 1 and 2) using different sampling periods to estimate symptomatic VE. Specifically, we began on day 20 (thus, a 20-day sampling period), and then repeated daily until the end of the epidemic. We conducted this analysis using both high and low default values of efficacy against susceptibility (0.55 and 0.1, respectively). We also varied the prevalence of etiologies of COVID-like symptoms to explore its influence on the magnitude of bias across the three testing scenarios.

We repeated the above analyses (with default parameters as per Table 1) under a range of epidemic and immunological properties to determine whether varying these properties could shape the magnitude of the bias (Objectives 2 and 3). First, we varied efficacy against susceptibility (0.1 to 0.9) and efficacy against infectiousness (0.1 to 0.9). We then varied the underlying epidemic potential - the probability of transmission (0.033 to 0.11^31–34)^.

Results for Objective 1 were reported for the entire sampling period (20 - 150 and 20 - 100 days for high and low efficacy against susceptibility, respectively); Results for Objectives 2 and 3 were reported at a single time point - the highest SARS-CoV-2 epidemic growth point (i.e. the time when the epidemic experienced its highest positive growth [inflection point]). A single time point was used for Objectives 2 and 3 so VE estimates could be easily compared across levels of epidemic and immunological properties.

All simulations were conducted using R^58^ (version 4.3.1) with computations performed on the Niagara supercomputer at the SciNet HPC Consortium^59,60^ (Supplementary 2: Computing Resources).

## Code Availability

The code to replicate all analyses is available on GitHub (https://github.com/mishra-lab/testing_bias.git).

## Data Availability

All simulated data produced for this study are available online on GitHub

https://github.com/mishra-lab/testing_bias.git

## Acknowledgements

This work was supported by the following: Canadian Institutes of Health Research (grant no. GA1-177697, VS1-175536; VS2-175581), the Natural Sciences and Engineering Research Council of Canada through the Mathematics for Public Health program (Emerging Infectious Disease Modeling grant no. RGPID-560523-2020), and the Data Sciences Institute and the Temerty Centre for AI Research and Education in Medicine at the University of Toronto by grant number DSI-CGY3R1P15.

SM is supported by a Tier 2 Canada Research Chair in Mathematical Modeling and Program Science award (grant no. CRC-950-232643). BS is supported by a Tier 1 Canada Research Chair in Economics of Infectious Diseases (grant No. CRC-2022-00362). JCK is supported by a Clinician-Scientist Award from the University of Toronto Department of Family and Community Medicine.

Computations were performed on the niagara supercomputer at the SciNet HPC Consortium. SciNet is funded by Innovation, Science and Economic Development Canada; the Digital Research Alliance of Canada; the Ministry of Colleges and Universities, Ontario; and the University of Toronto.

We thank Dr. Jesse Knight, Huiting Ma, Siyi Wang, and Dr. Alexander Whitlock, for helpful discussions during research group meetings and in preparation for scientific presentations, as well as Dr.Susana Monge and her team for their feedback on our methods and model results. As always, KB would like to thank Cedric B. Hunter and Winston Irwin for their continued support.

