## Supplementary for "Impact of unequal testing on vaccine effectiveness estimates across two study designs: a simulation study"

### Supplementary 1: Results

Timing of Testing

We compared the average time until testing for vaccinated and unvaccinated individuals using our three testing scenarios: equal testing, moderately unequal testing, and highly unequal testing. We simulated a population of 100,000 symptomatic individuals whereby 50% of individuals were vaccinated and 50% were unvaccinated. See Supplementary 2: Testing and Healthcare Engagement for details on how testing scenarios were implemented.

We found that with equal testing by vaccination status the average time until testing was the same for vaccinated and unvaccinated individuals (Table S1). When testing was unequal, unvaccinated individuals experienced a longer average time until testing than vaccinated individuals (Table S1). The difference in this average time grew with the degree of testing differences. When testing differences were moderately unequal, unvaccinated individuals tested 0.35 days later than unvaccinated individuals (4.55 vs 4.20); when testing was highly unequal by vaccination status, unvaccinated individuals tested 0.62 days later than unvaccinated (4.83 vs. 4.21).

**Table S1:** Average day of first test by vaccination status across our three testing scenarios.

| | **Average Day of First Test** |
|---|---|
| *Equal Testing Scenario* | |
| Vaccinated | 4.19 |
| Unvaccinated | 4.19 |
| *Moderately Unequal Testing Scenario* | |
| Vaccinated | 4.20 |
| Unvaccinated | 4.55 |
| *Highly Unequal Testing Scenario* | |
| Vaccinated | 4.21 |
| Unvaccinated | 4.83 |

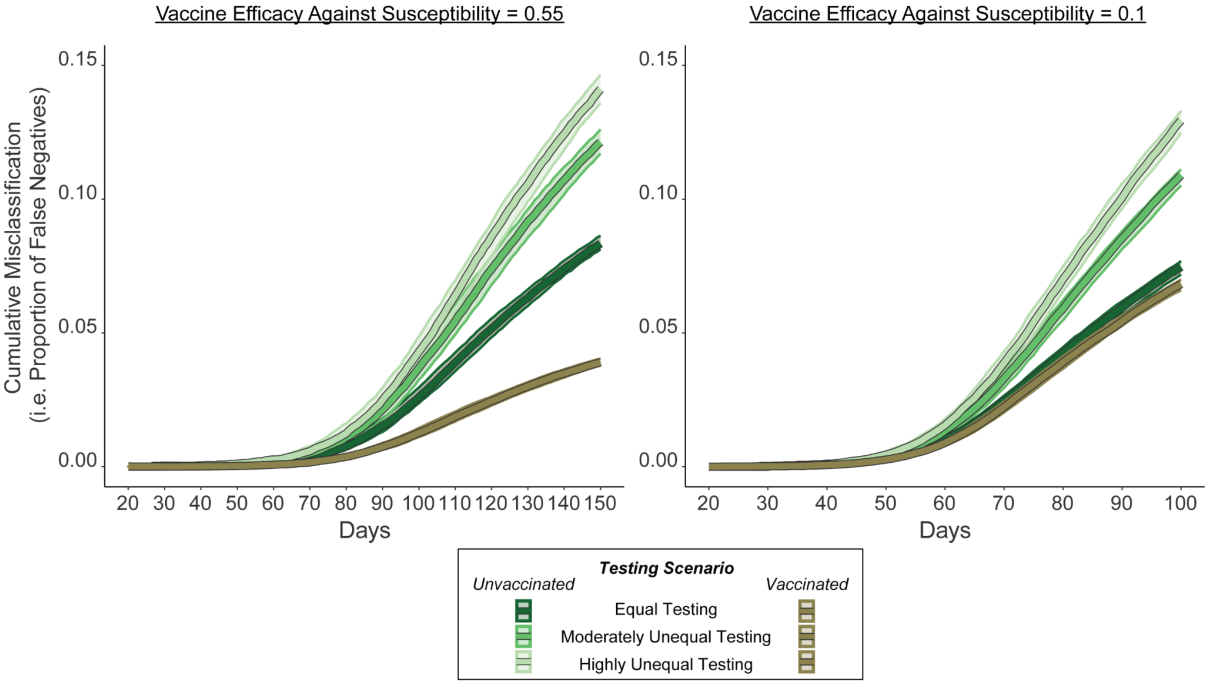


**Supplementary Fig. 1:** The cumulative proportion of misclassifications (i.e. false negatives) by vaccination status for the test negative design over time. Testing scenario depicts the degree of cumulative misclassification (i.e. cumulative proportion of false negatives) given equal testing, moderately unequal testing (vaccinated with 1.76x higher testing), and highly unequal testing (vaccinated with 2.36x higher testing) for unvaccinated individuals (green) and vaccinated individuals (brown). Lines depict the median value of 100 epidemic realizations and the shaded area represents the interquartile range.

### Misclassification Example

An individual can be misclassified in the test-negative design when they have had a prior symptomatic SARS-CoV-2 infection during the sampling period but were only tested when they had COVID-like symptoms from other etiologies. For example, an individual is infected with symptomatic SARS-CoV-2 early in the epidemic and by chance does not receive testing. That individual is not included in the sample for the test-negative design at this point in time. At a later point in time, the same individual has COVID-like symptoms due to other etiologies and by chance, receives testing (Supplementary Fig. 2). The individual is then classified as negative during this sampling period even though they had a symptomatic SARS-CoV-2 infection (i.e. a misclassification). Note the chances of this misclassification increases when there is a higher probability of having COVID-like symptoms from other etiologies (Supplementary Fig. 3).

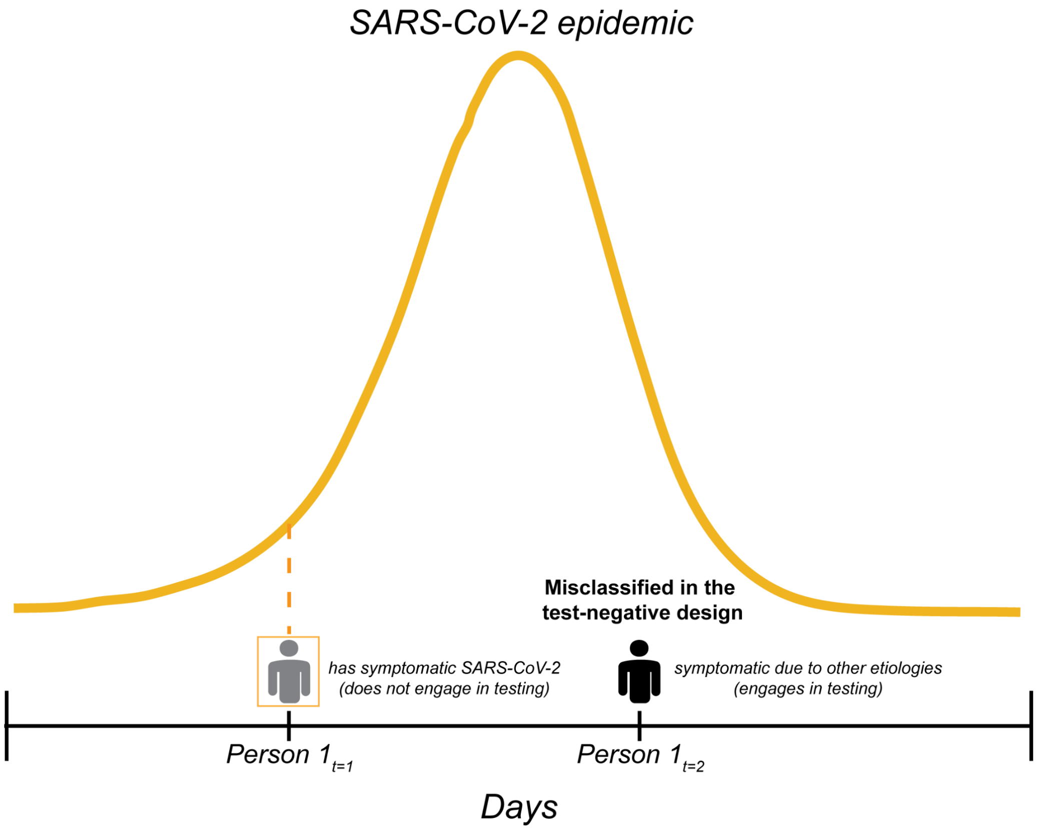


**Supplementary Fig. 2** Example of a misclassification in the test-negative design using an individual (person 1) at two different timepoints ($t$=1 and $t$=2). At $t$=1, the individual is infected with symptomatic SARS-CoV-2 and does not engage in testing. At $t$=2, the individual has COVID-like symptoms due to other etiologies and engages in testing

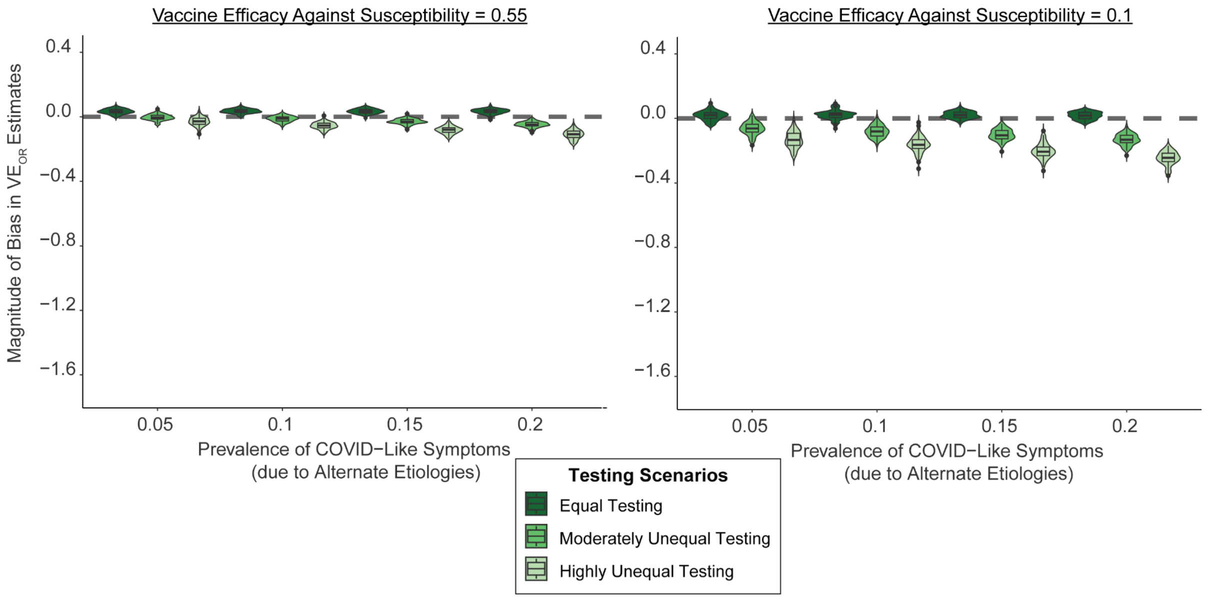


**Supplementary Fig. 3:** Magnitude of bias in symptomatic vaccine effectiveness (symptomatic VE) estimates from the test-negative design ( $VE_{OR}$) across testing scenarios and prevalences of COVID-like symptoms (due to alternate etiologies). Symptomatic VE estimates were measured across both higher and lower levels of vaccine efficacy against susceptibility (0.55 and 0.1), which reflects the true symptomatic VE (grey dashed line). Each epidemic scenario was simulated 100 times with symptomatic VE estimates calculated at the point of the highest positive epidemic growth per SARS-CoV-2 epidemic. The median time of sampling was $t$ = 50 for 0.1 and $t$ = 73 for 0.55.

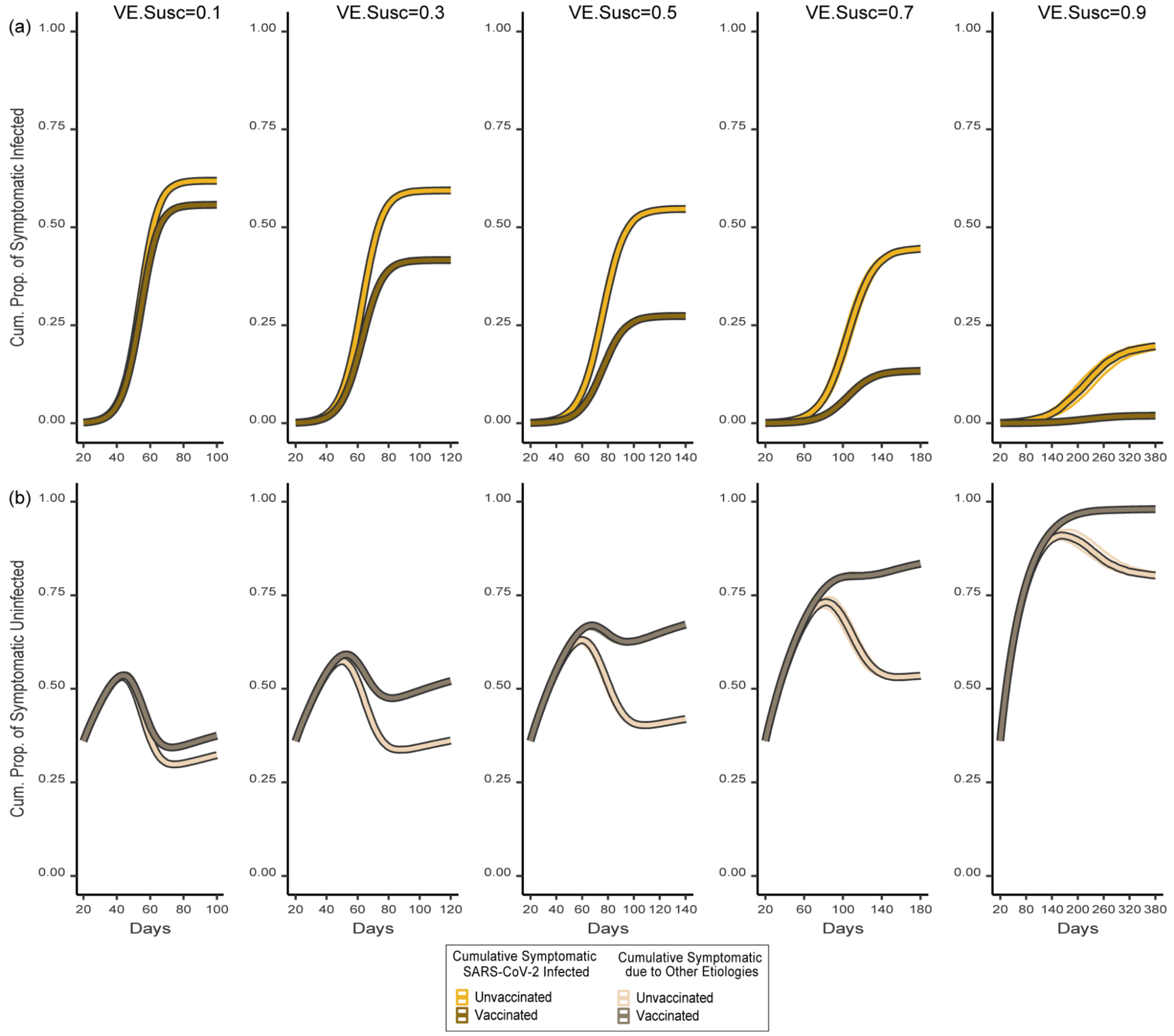


**Supplementary Fig. 4:** Cumulative proportion of symptomatic SARS-CoV-2 individuals (a) and cumulative proportion of symptomatic individuals due to other etiologies (b) over time and across levels of vaccine efficacy against susceptibility (VE.Susc). The cumulative proportion of symptomatic individuals due to other etiologies includes only those individuals with no prior symptomatic SARS-CoV-2 infection. Line colour represents vaccination status and SARS-CoV-2 symptomatic infection status (vaccinated and have/had SARS-CoV-2 infection [dark gold]; unvaccinated and have/had SARS-CoV-2 infection [yellow]; vaccinated symptomatic but never infected with SARS-CoV-2 [dark brown]; unvaccinated symptomatic but never infected with SARS-CoV-2 [beige]); Line values depict the median value of 100 epidemic realizations with shaded regions representing the interquartile range. Figure panel columns represent different levels of vaccine efficacy against susceptibility. Note that the x-axis scale changes by column with higher levels of efficacy against susceptibility representing a longer period of time.

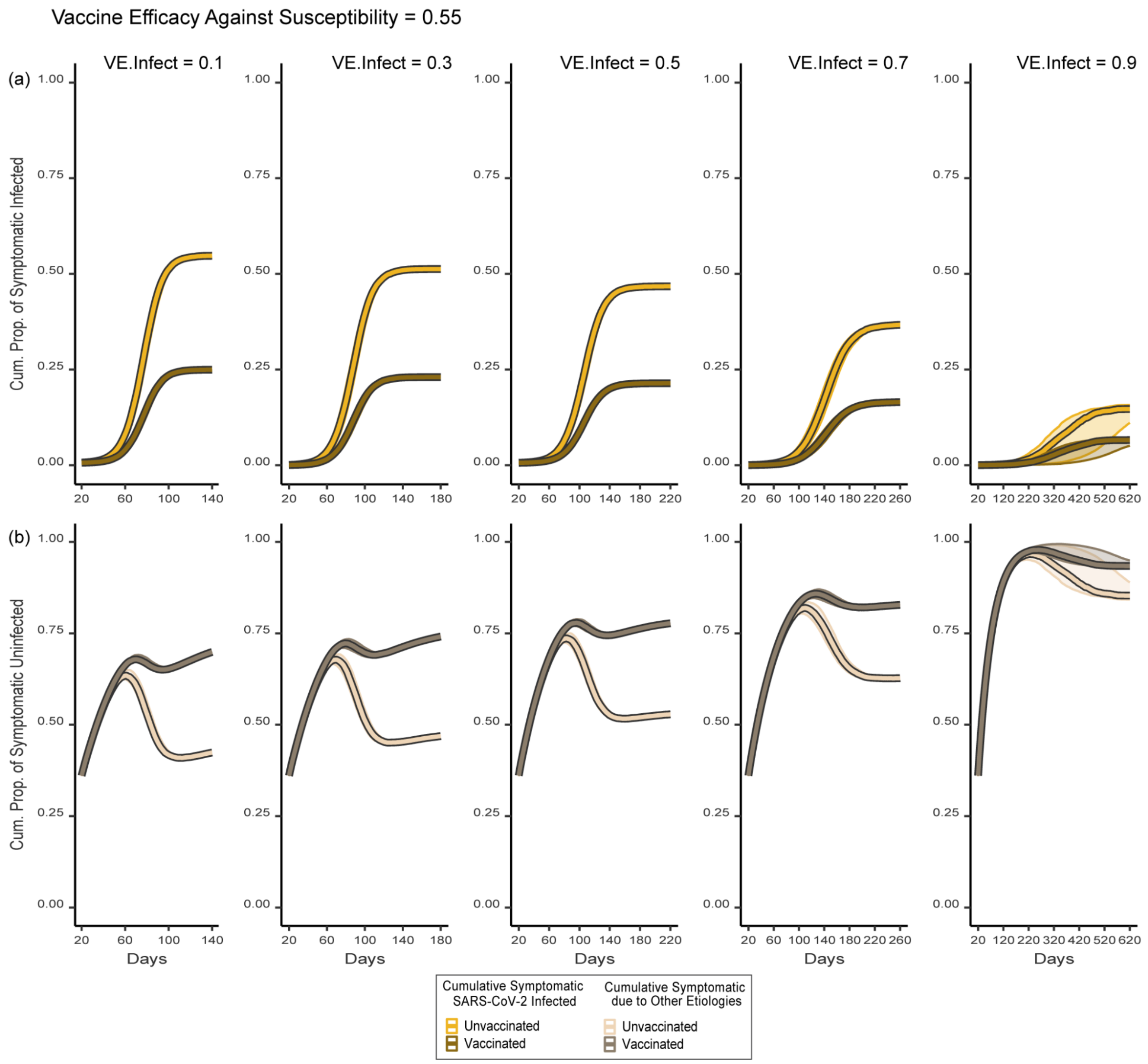


**Supplementary Fig. 5:** Cumulative proportion of symptomatic SARS-CoV-2 individuals (a) and cumulative proportion of symptomatic individuals due to other etiologies (b) over time and across levels of vaccine efficacy against infectiousness (VE.Infect), assuming a higher vaccine efficacy against susceptibility (0.55). The cumulative proportion of symptomatic individuals due to other etiologies includes only those individuals with no prior symptomatic SARS-CoV-2 infection. Line colour represents vaccination status and SARS-CoV-2 infection status (vaccinated and have/had SARS-CoV-2 infection [dark gold]; unvaccinated and have/had SARS-CoV-2 infection [yellow]; vaccinated symptomatic but never infected with SARS-CoV-2 [dark brown]; unvaccinated symptomatic but never infected with SARS-CoV-2 [beige]); Line values depict the median value of 100 epidemic realizations with shaded regions representing the interquartile range. Figure panel columns represent different levels of vaccine efficacy against infectiousness. Note that the x-axis scale changes by column with higher levels of efficacy against infectiousness representing a longer period of time.

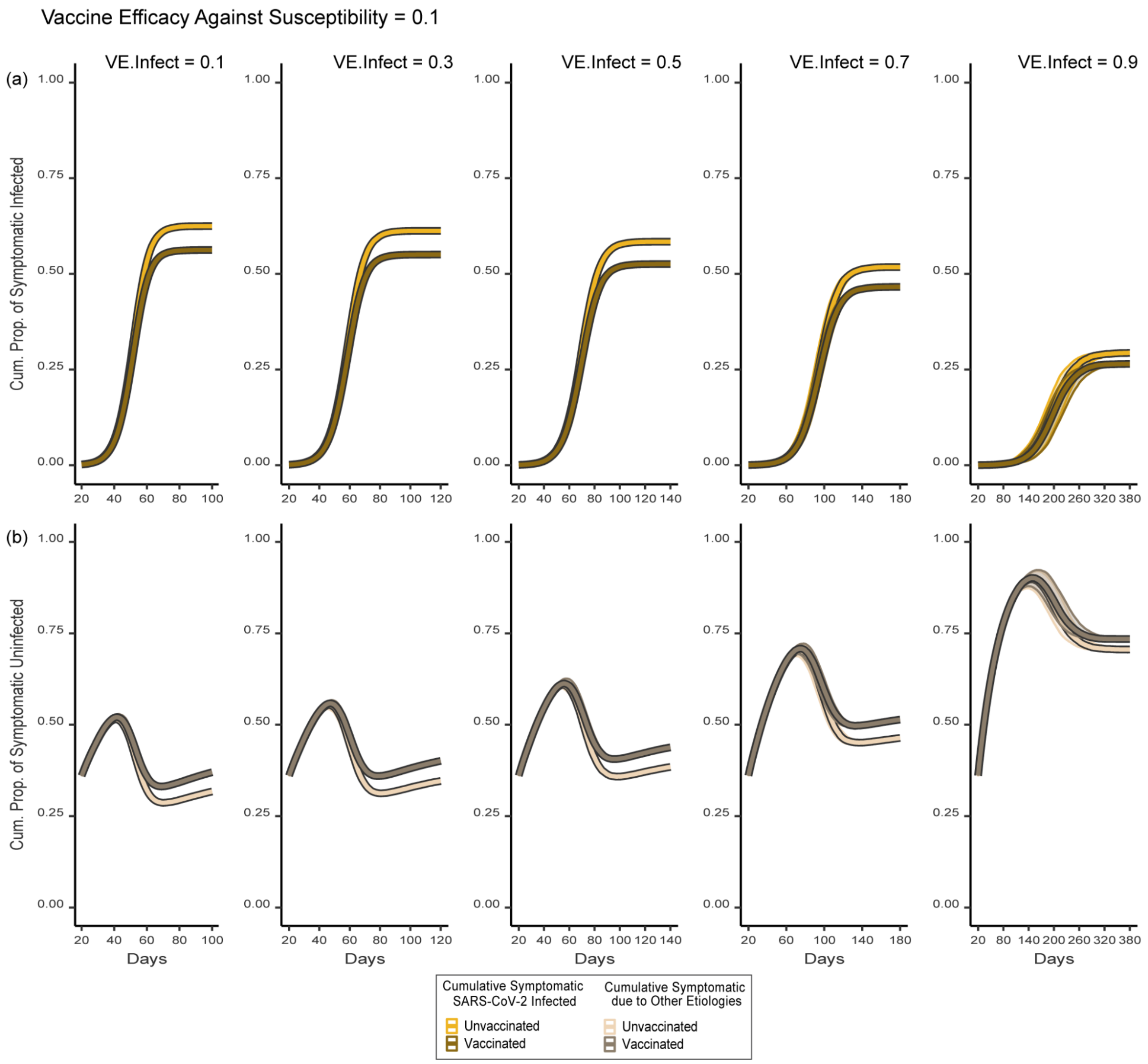


**Supplementary Fig. 6:** Cumulative proportion of symptomatic SARS-CoV-2 individuals (a) and cumulative proportion of symptomatic individuals due to other etiologies (b) over time and across levels of vaccine efficacy against infectiousness (VE.Infect), assuming a lower vaccine efficacy against susceptibility (0.1). The cumulative proportion of symptomatic individuals due to other etiologies includes only those individuals with no prior symptomatic SARS-CoV-2 infection. Line colour represents vaccination status and SARS-CoV-2 infection status (vaccinated and have/had SARS-CoV-2 infection [dark gold]; unvaccinated and have/had SARS-CoV-2 infection [yellow]; vaccinated symptomatic but never infected with SARS-CoV-2 [dark brown]; unvaccinated symptomatic but never infected with SARS-CoV-2 [beige]); Line values depict the median value of 100 epidemic realizations with shaded regions representing the interquartile range. Figure panel columns represent different levels of vaccine efficacy against infectiousness. Note that the x-axis scale changes by column with higher levels of efficacy against infectiousness representing a longer period of time.

Moderating Effect Calculations

The consequences of changing vaccine efficacies on the calculations of the relative risk and odds ratios explain the differences in the moderating effects on the testing bias by study design.

*Vaccine Efficacy Against Susceptibility*

With larger efficacy against susceptibility and unequal testing, the growing absolute difference between symptomatic infections by vaccination status meant that the ratio of the numerators in the relative risk equation ($\frac{CI_V}{CI_U}$; equation 1) became smaller (Supplementary Fig. 7). As a result, as efficacy against susceptibility increased, the cohort design led to a reduction of the bias.

At the same time, when testing was unequal, the ratio of the denominators ($\frac{CU_V}{CU_U}$; equation 2) of the odds ratio calculation also became smaller. However, the change in the ratio of the denominators was smaller than the change in the ratio of the numerators (Supplementary Fig. 8). As a result, when testing was unequal, the test-negative design led to a smaller reduction of bias (because both the numerator and denominators of the ratio were affected) compared to the cohort design (that was influenced only on the numerators) .

*Vaccine Efficacy Against Infectiousness*

Similar to efficacy against susceptibility, larger efficacy against infectiousness and unequal testing, caused the ratio of the numerators in the relative risk equation ($\frac{CI_V}{CI_U}$; equation 1) to become smaller (Supplementary Fig. 9). However, due to the more proportionate reductions in cumulative infections by vaccination status caused by increasing efficacy against infectiousness (Supplementary Fig. 6), this change was substantially smaller than the change observed with efficacy against susceptibility (Supplementary Fig. 7). This more proportionate change led to only small, minimal changes in the magnitude of the bias as efficacy against infectiousness increased with the cohort design.

At the same time, with unequal testing, the ratio of the denominators ($\frac{CU_V}{CU_U}$; equation 2) of the odds ratio calculation became smaller as efficacy against infectiousness increased. In contrast to efficacy against susceptibility, the change in the ratio of the denominators was greater than the change in the ratio of the numerators (Supplementary Fig. 10). Therefore, the magnitude of the bias increased with larger efficacy against infectiousness with the test-negative design.

*Epidemic Potential (Probability of Transmission)*

Changing epidemic potential altered epidemic dynamics (Supplementary Fig. 11-12), which also affected the relative risk and odds ratio calculations.With smaller epidemic potential (i.e. lower probability of transmission), and unequal testing, the ratio of the numerators and the denominators followed similar patterns as with larger efficacy against infectiousness (Supplementary Fig. 13-14). The resulting effect was the magnitude of the bias minimally decreasing with the cohort design and increasing more with the test-negative design with decreasing epidemic potential.  These patterns emerge because, similar to increasing efficacy against infectiousness, reductions in the epidemic potential led to more proportionate reductions in cumulative infections (Supplementary Fig. 12).

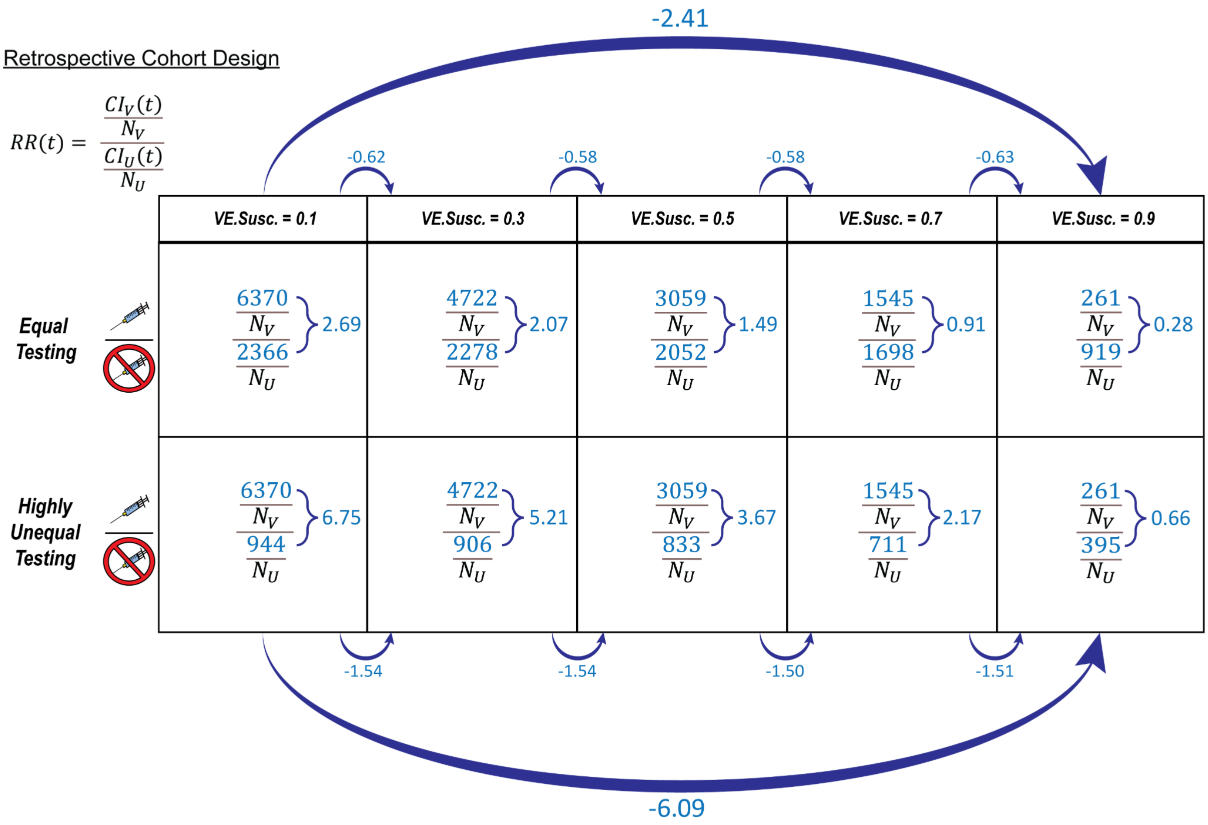


**Supplementary Fig. 7:** Changes in the ratio of the numerators in the retrospective cohort design's relative risk equation $RR(t)$) across levels of vaccine efficacy against susceptibility and testing scenarios. $CI_V\ (t)$ and $CI_U\ (t)$ are the cumulative numbers of symptomatic infected for vaccinated and unvaccinated that have been tested and diagnosed (tested positive) at time *t,* respectively; and $N_V$ and $N_U$ are the total numbers of vaccinated and unvaccinated individuals, respectively. Rows represent testing scenarios: equal testing (equal testing by vaccination status) (top); and highly unequal testing (vaccinated have 2.36 higher testing than unvaccinated) (bottom); columns represent level of vaccine efficacy against susceptibility. The blue arrows represent the difference between ratios of the numerators (i.e. $CI_V\ (t)$ over $CI_U\ (t)$) from one cell to another. All numbers are the median value of 100 epidemic realizations taken at the highest SARS-CoV-2 epidemic growth point (i.e. the time when the epidemic experienced its highest positive growth) for each scenario.

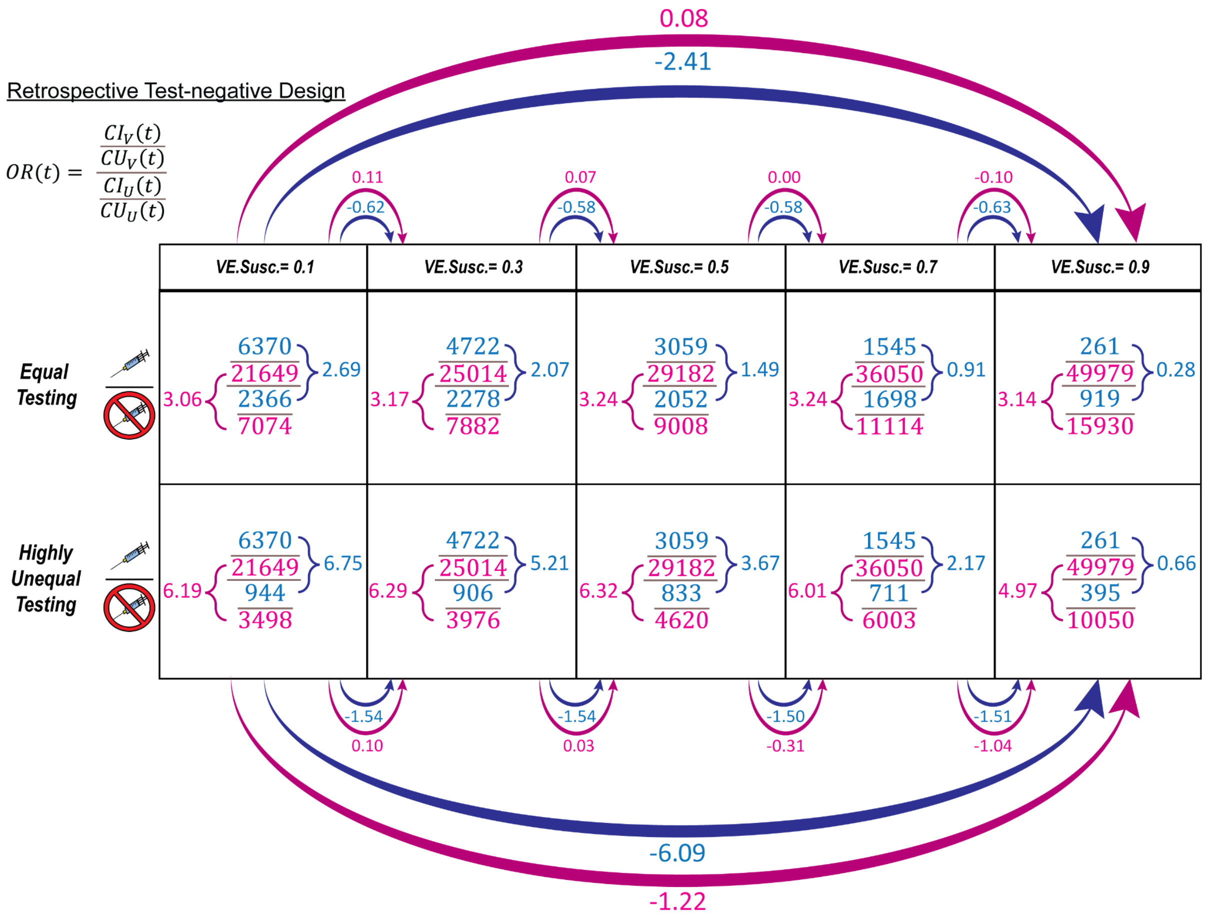


**Supplementary Fig. 8:** Changes in the ratio of the numerators and the ratio of the denominators in the retrospective test-negative design's odds ratio equation ($OR(t)$) across levels of vaccine efficacy against susceptibility and testing scenarios. $CI_V\ (t)$ and $CI_U\ (t)$ are the cumulative numbers of symptomatic infected for vaccinated and unvaccinated that have been tested and diagnosed (tested positive) at time *t*, respectively; $CU_V\ (t)$ and $CU_U\ (t)$ are the cumulative numbers of symptomatic individuals (with COVID-like symptoms) for vaccinated and unvaccinated populations who tested negative for SARS-COV-2 and had no prior positive tests for SARS-COV-2 by time *t*, respectively. Rows represent testing scenarios: equal testing (equal testing by vaccination status) (top); and highly unequal testing (vaccinated have 2.36 higher testing than unvaccinated) (bottom); columns represent level of vaccine efficacy against susceptibility. The blue arrows represent the difference between the ratios of the numerators (i.e. $\frac{CI_V(t)}{CI_U(t)}$) from one cell to another; the pink arrows represent the difference between the ratio of the denominators (i.e. $\frac{CU_V\ (t)}{CU_U\ (t)}$). All numbers are the median value of 100 epidemic realizations taken at the highest SARS-CoV-2 epidemic growth point (i.e. the time when the epidemic experienced its highest positive growth) for each scenario.

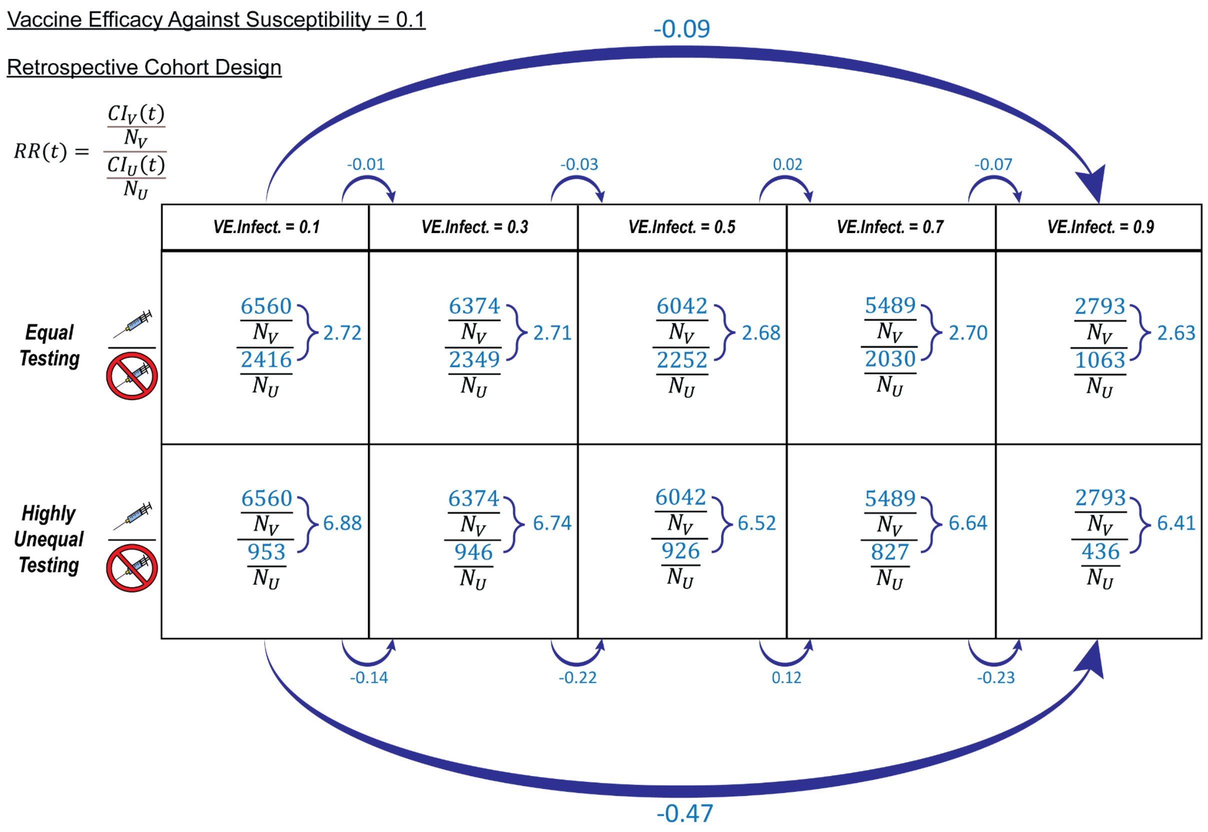


**Supplementary Fig. 9:** Changes in the ratio of the numerators in the retrospective cohort design's relative risk equation $RR(t)$) across levels of vaccine efficacy against infectiousness and testing scenarios given a low vaccine efficacy against susceptibility (0.1). $CI_V\ (t)$ and $CI_U\ (t)$ are the cumulative numbers of symptomatic infected for vaccinated and unvaccinated that have been tested and diagnosed (tested positive) at time *t*, respectively; and $N_V$ and $N_U$ are the total numbers of vaccinated and unvaccinated individuals, respectively. Rows represent testing scenarios: Equal Testing (equal testing by vaccination status) (top); and Highly Unequal Testing (vaccinated have 2.36 higher testing than unvaccinated) (bottom); columns represent level of vaccine efficacy against infectiousness. The blue arrows represent the difference between ratios of the numerators (i.e. $CI_V\ (t)$ over $CI_U\ (t)$) from one cell to another. All numbers are the median value of 100 epidemic realizations taken at the highest SARS-CoV-2 epidemic growth point (i.e. the time when the epidemic experienced its highest positive growth) for each scenario.

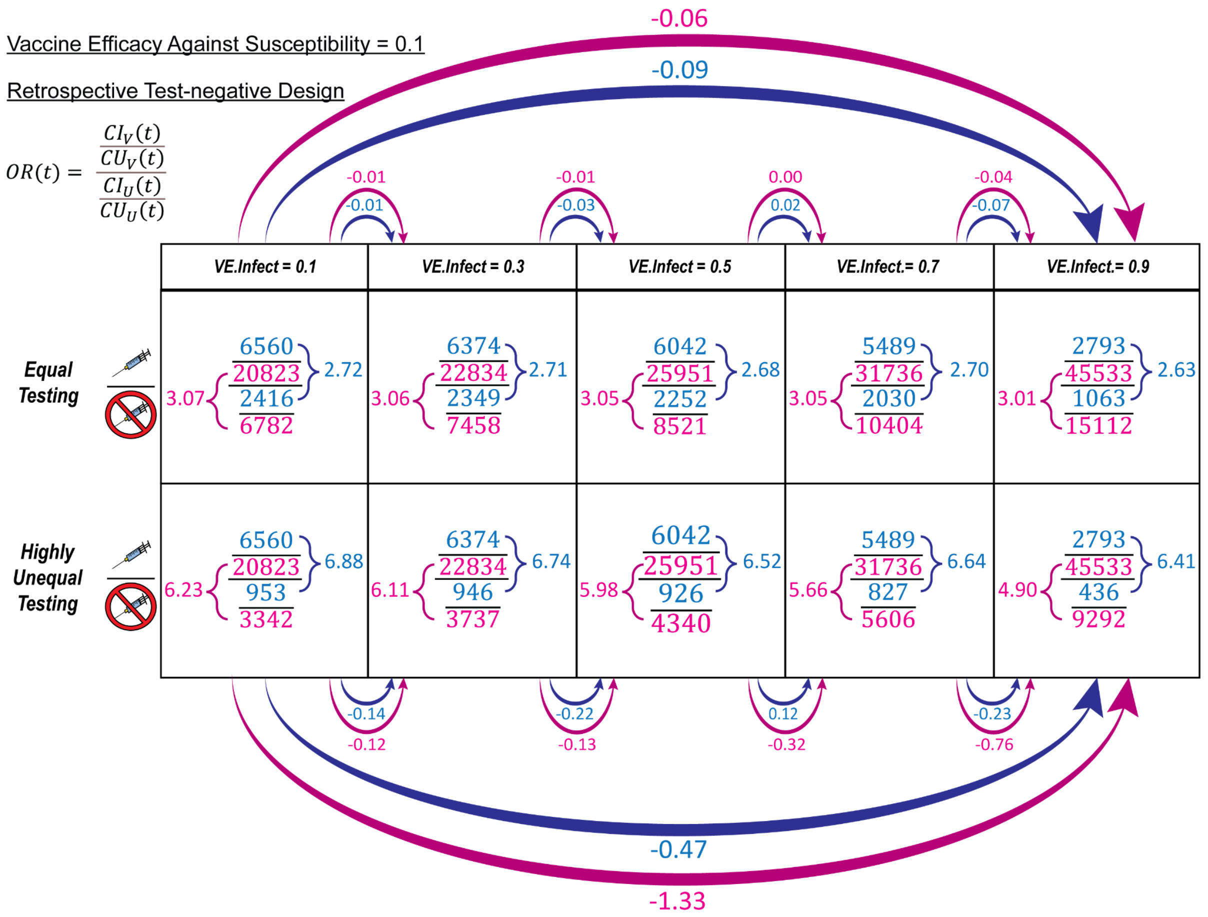


**Supplementary Fig. 10:** Changes in the ratio of the numerators and the ratio of the denominators in the retrospective test-negative design's odds ratio equation ($OR(t)$) across levels of vaccine efficacy against infectiousness and testing scenarios given a low vaccine efficacy against susceptibility (0.1). $CI_V\ (t)$ and $CI_U\ (t)$ are the cumulative numbers of symptomatic infected for vaccinated and unvaccinated that have been tested and diagnosed (tested positive) at time *t,* respectively; $CU_V\ (t)$ and $CU_U\ (t)$ are the cumulative numbers of symptomatic individuals (with COVID-like symptoms) for vaccinated and unvaccinated populations who tested negative for SARS-COV-2 and had no prior positive tests for SARS-COV-2 by time *t,* respectively. Rows represent testing scenarios: Equal Testing (equal testing by vaccination status) (top); and Highly Unequal Testing (vaccinated have 2.36 higher testing than unvaccinated) (bottom); columns represent level of vaccine efficacy against infectiousness. The blue arrows represent the difference between the ratios of the numerators (i.e. $\frac{CI_V(t)}{CI_U(t)}$) from one cell to another; the pink arrows represent the difference between the ratio of the denominators (i.e. $\frac{CU_V\ (t)}{CU_U\ (t)}$). All numbers are the median value of 100 epidemic realizations taken at the highest SARS-CoV-2 epidemic growth point (i.e. the time when the epidemic experienced its highest positive growth) for each scenario.

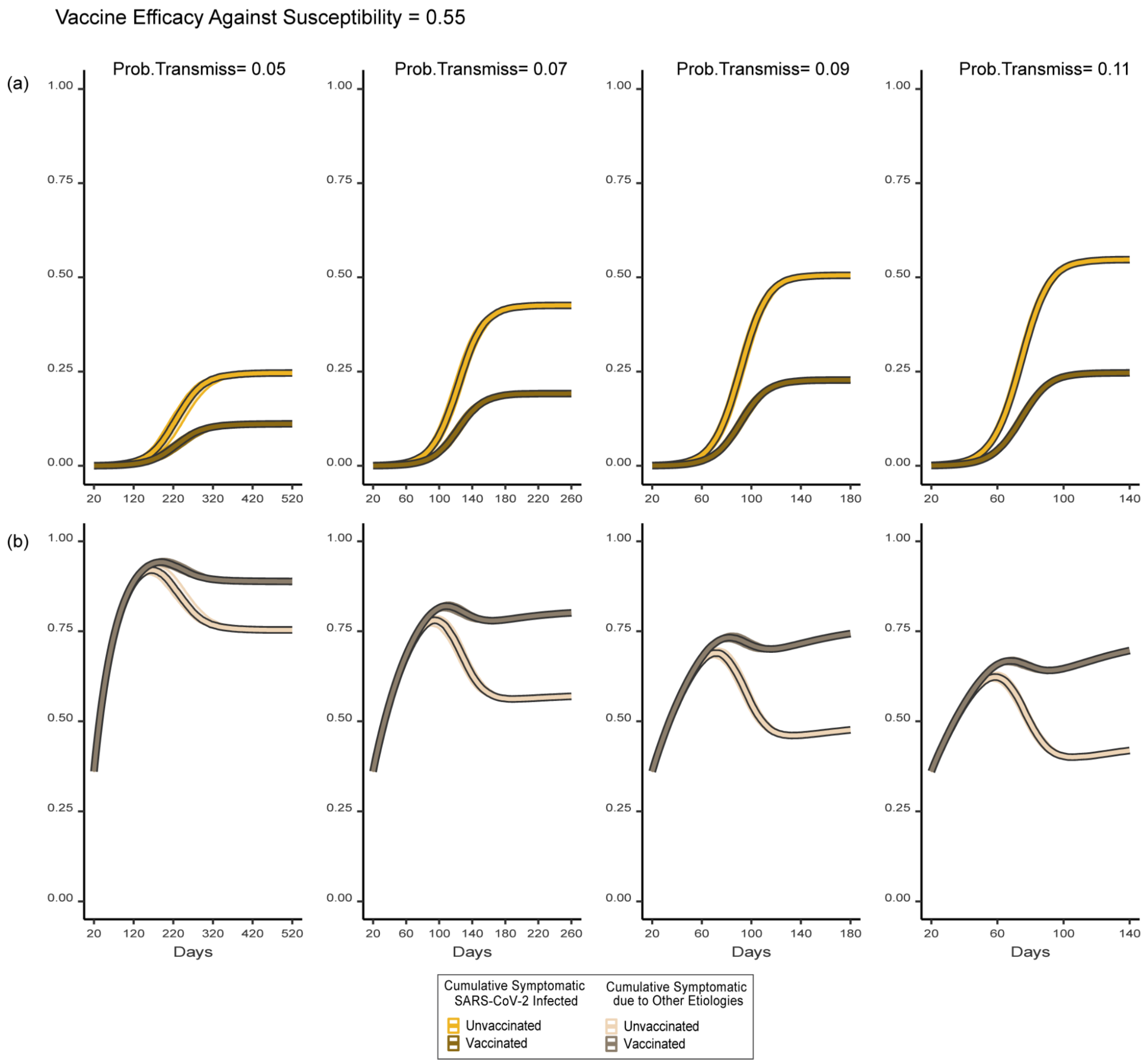


**Supplementary Fig. 11:** Cumulative proportion of symptomatic SARS-CoV-2 individuals (a) and cumulative proportion of symptomatic individuals due to other etiologies (b) over time and across probabilities of transmission (Prob.Transmiss), assuming a higher vaccine efficacy against susceptibility (0.55). The cumulative proportion of symptomatic individuals due to other etiologies includes only those individuals with no prior symptomatic SARS-CoV-2 infection. Line colour represents vaccination status and SARS-CoV-2 infection status (vaccinated and have/had SARS-CoV-2 infection [dark gold]; unvaccinated and have/had SARS-CoV-2 infection [yellow]; vaccinated symptomatic but never infected with SARS-CoV-2 [dark brown]; unvaccinated symptomatic but never infected with SARS-CoV-2 [beige]); Line values depict the median value of 100 epidemic realizations with shaded regions representing the interquartile range. Figure panel columns represent different levels of transmission. Note that the x-axis scale changes by column with lower probability of transmission representing a longer period of time.

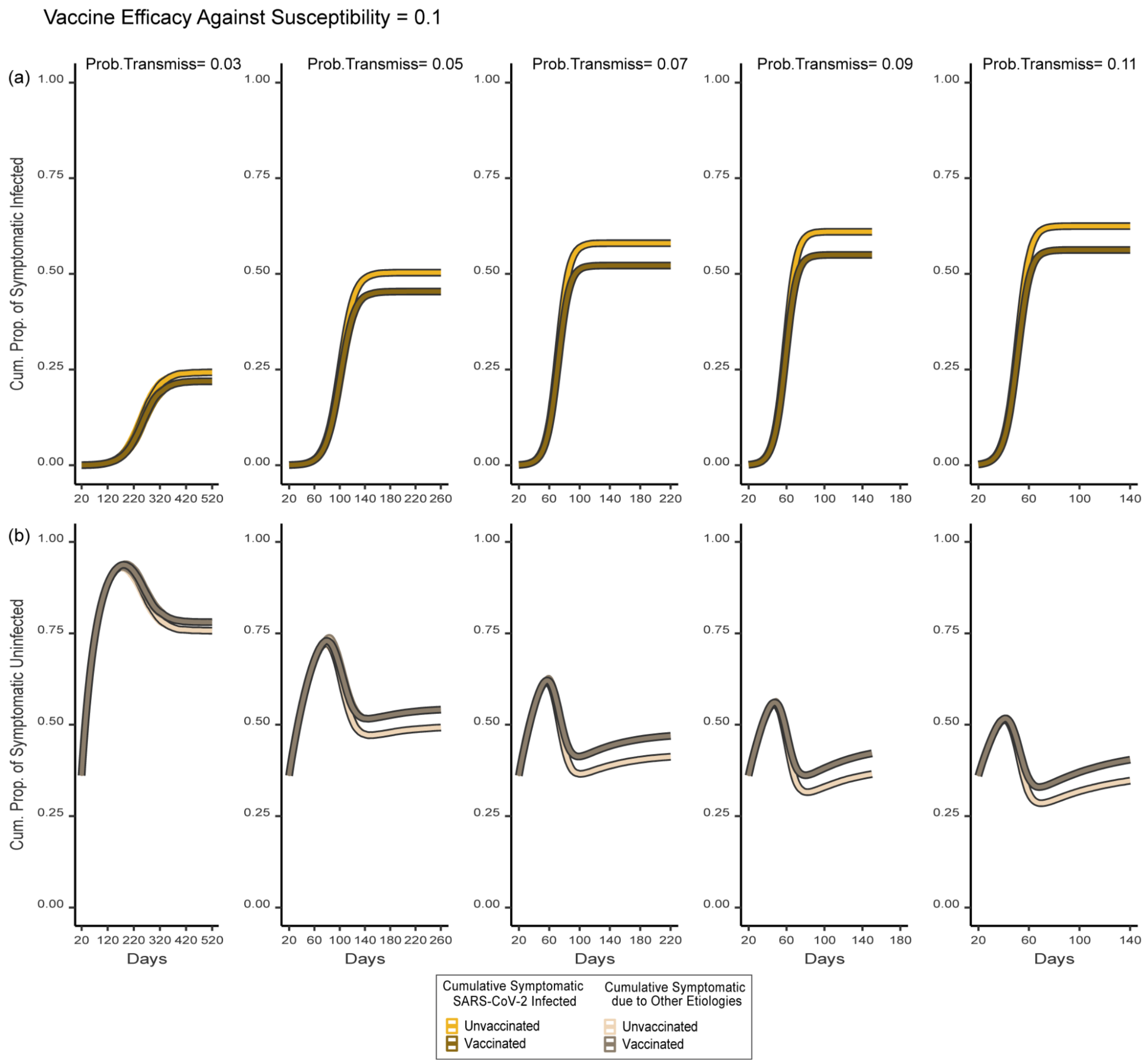


**Supplementary Fig. 12:** Cumulative proportion of symptomatic SARS-CoV-2 individuals (a) and cumulative proportion of symptomatic individuals due to other etiologies (b) over time and across probabilities of transmission (Prob.Transmiss), assuming a lower vaccine efficacy against susceptibility (0.1). The cumulative proportion of symptomatic individuals due to other etiologies includes only those individuals with no prior symptomatic SARS-CoV-2 infection. Line colour represents vaccination status and SARS-CoV-2 infection status (vaccinated and have/had SARS-CoV-2 infection [dark gold]; unvaccinated and have/had SARS-CoV-2 infection [yellow]; vaccinated symptomatic but never infected with SARS-CoV-2 [dark brown]; unvaccinated symptomatic but never infected with SARS-CoV-2 [beige]); Line values depict the median value of 100 epidemic realizations with shaded regions representing the interquartile range. Figure panel columns represent different levels of transmission. Note that the x-axis scale changes by column with lower probability of transmission representing a longer period of time.

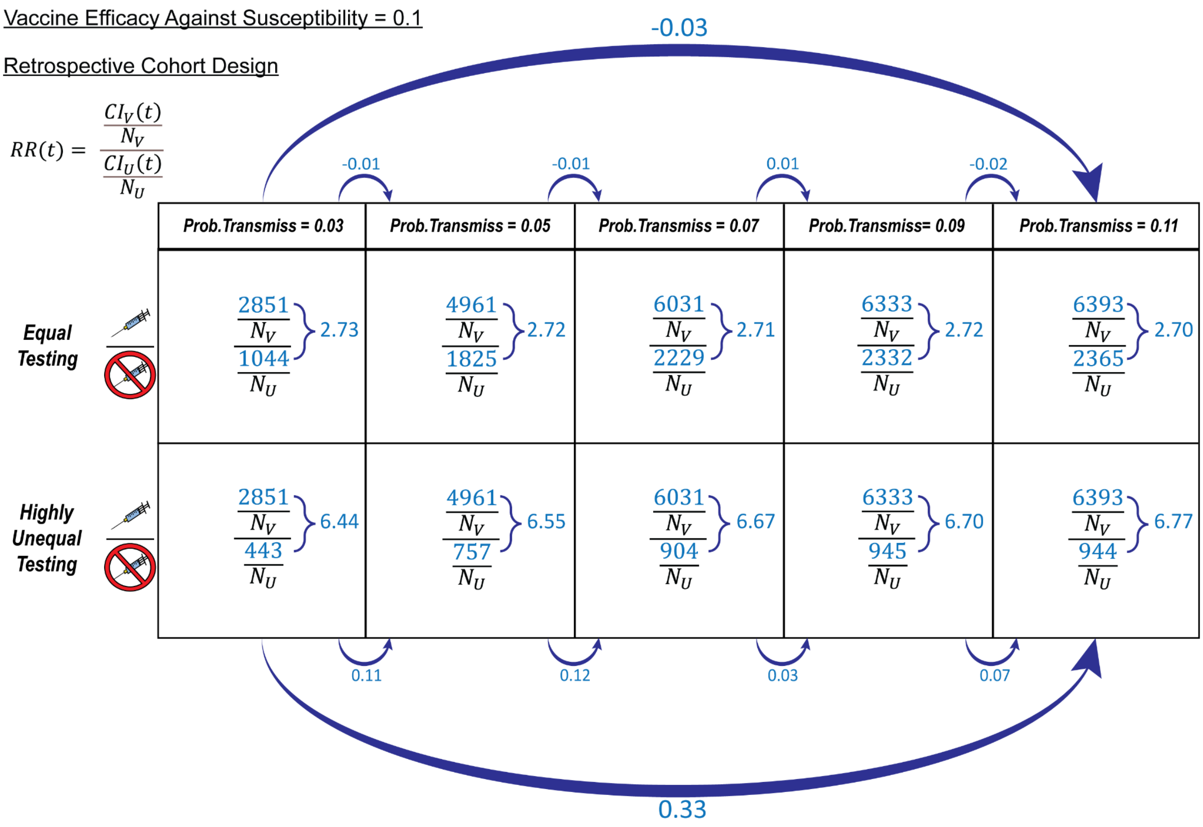


**Supplementary Fig. 13:** Changes in the ratio of the numerators in the retrospective cohort design's relative risk equation $RR(t)$) across probabilities of transmission (Prob. Transmiss) and testing scenarios given a low vaccine efficacy against susceptibility (0.1). $CI_V\ (t)$ and $CI_U\ (t)$ are the cumulative numbers of symptomatic infected for vaccinated and unvaccinated that have been tested and diagnosed (tested positive) at time *t*, respectively; and $N_V$ and $N_U$ are the total numbers of vaccinated and unvaccinated individuals, respectively. Rows represent testing scenarios: Equal Testing (equal testing by vaccination status) (top); and Highly Unequal Testing (vaccinated have 2.36 higher testing than unvaccinated) (bottom); columns represent level of transmission. The blue arrows represent the difference between ratios of the numerators (i.e. $CI_V\ (t)$ over $CI_U\ (t)$) from one cell to another. All numbers are the median value of 100 epidemic realizations taken at the highest SARS-CoV-2 epidemic growth point (i.e. the time when the epidemic experienced its highest positive growth) for each scenario.

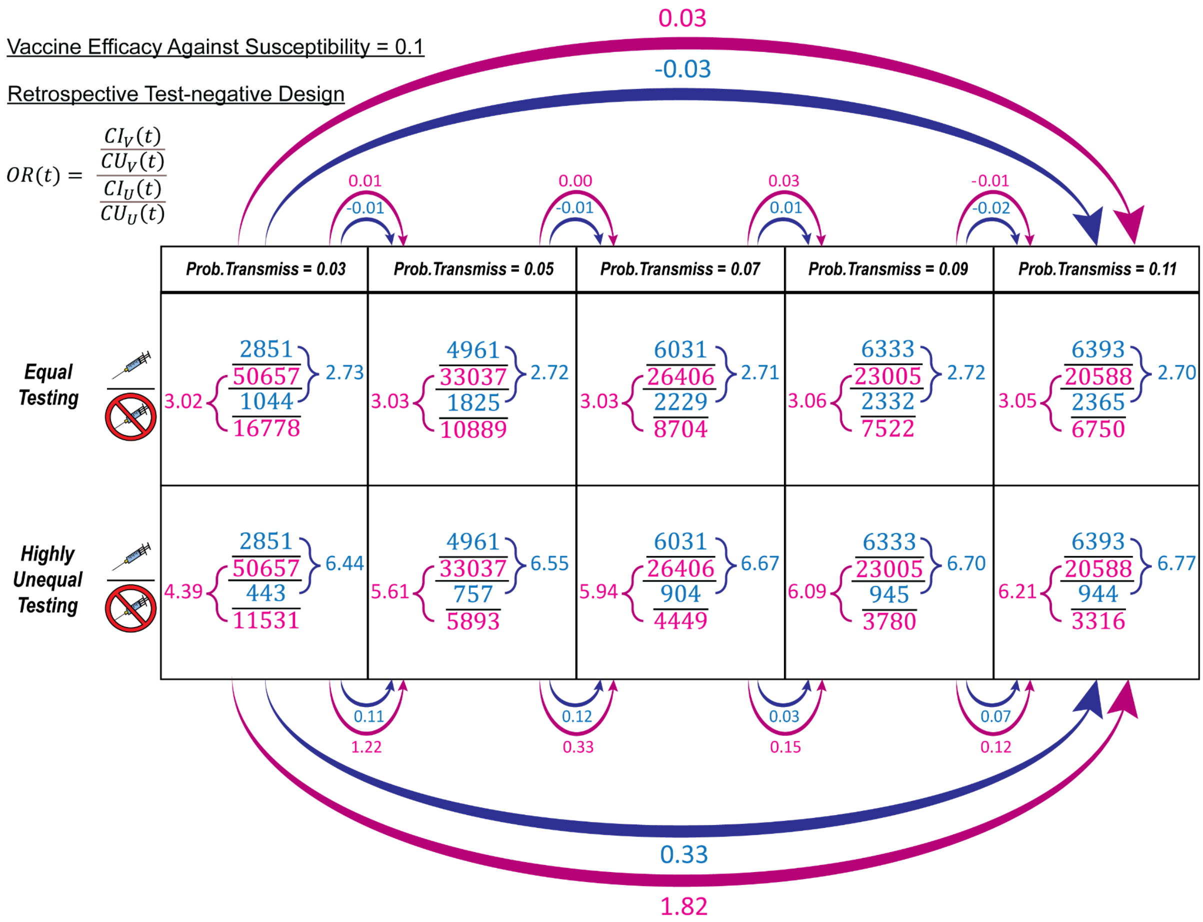


**Supplementary Fig. 14:** Changes in the ratio of the numerators and the ratio of the denominators in the retrospective test-negative design's odds ratio equation ($OR(t)$) across probabilities of transmission (Prob. Transmiss) and testing scenarios given a low vaccine efficacy against susceptibility (0.1). $CI_V\ (t)$ and $CI_U\ (t)$ are the cumulative numbers of symptomatic infected for vaccinated and unvaccinated that have been tested and diagnosed (tested positive) at time *t*, respectively; $CU_V\ (t)$ and $CU_U\ (t)$ are the cumulative numbers of symptomatic individuals (with COVID-like symptoms) for vaccinated and unvaccinated populations who tested negative for SARS-COV-2 and had no prior positive tests for SARS-COV-2 by time *t*, respectively. Rows represent testing scenarios: Equal Testing (equal testing by vaccination status) (top); and Highly Unequal Testing (vaccinated have 2.36 higher testing than unvaccinated) (bottom); columns represent levels of transmission. The blue arrows represent the difference between the ratios of the numerators (i.e. $\frac{CI_V(t)}{CI_U(t)}$) from one cell to another; the pink arrows represent the difference between the ratio of the denominators (i.e. $\frac{CU_V\ (t)}{CU_U\ (t)}$). All numbers are the median value of 100 epidemic realizations taken at the highest SARS-CoV-2 epidemic growth point (i.e. the time when the epidemic experienced its highest positive growth) for each scenario.

### Supplementary 2: Methods

#### Selection Bias

Test negative designs that restrict the study sample only to individuals who received a test may introduce a selection bias via selecting on a collider (also referred to as collider bias[1,2]. Testing is a collider because the exposure (vaccination) and the outcome (infection) both affect the likelihood of being tested (a collider is a variable that is influenced by two other variables[3]).

In studies that measure vaccine effectiveness against symptomatic infection, the study sample is often restricted to individuals who are symptomatic and testing for SARS-CoV-2 (depicted by the two "conditioned upon" circles in Fig. 1). When both symptomatic and testing for SARS-CoV-2 are conditioned upon, a selection bias can still be generated due to selecting/conditioning on a collider (Fig. 1B). Testing remains a collider as both vaccination and infection are associated with testing. While conditioning on symptomatic blocks the association between SARS-CoV-2 infection and testing (as symptomatic is a mediator), it also induces an association between SARS-CoV-2 infection and etiologies of COVID-like symptoms ("e" in Fig. 1B). If individuals who are not symptomatic do not have etiologies of COVID-like symptoms (an assumption in our study), a relationship exists between etiologies of COVID-like symptoms and testing for SARS-CoV-2 ("d" in Fig. 1). When both "d" and "e" exist, there is a pathway between infection and testing through which the selection bias can persist.

#### Contact Patterns

The network was generated using an Erdos Renyi random model assuming an average of 6 contacts. As the network is sparse - an average of 6 contacts with 100,000 individuals -, the degree distribution (i.e. distribution of contacts) for the Erdos Renyi random model approximates a Poisson distribution[4].

To ensure all individuals were connected to the main network, disconnected individual nodes (representing individuals) were randomly assigned a single connection to a node within the giant component (i.e. the connected portion of the network that contains a significant number of nodes). If there was a group of disconnected nodes, we would randomly select a single node from that group to connect to a node within the giant component. This modification to the network resulted in a negligible effect on the networks with the average degree remaining 6 for each network (rounding to the hundredth decimal).

#### SARS-COV-2 Transmission Dynamics

Each individual (agent) in the model contains an internal "SEIR state" that provides their current SARS-CoV-2 state at any given time. Daily contact between individuals was pre-determined by the contact network. The probability of individuals becoming infected with SARS-CoV-2 and the probability of developing symptoms were both implemented using draws from Bernoulii distributions:

$f(x;p) = p^x(1-p)^{1-x}$ for $x \in \{0,1\}$

where p is the probability of becoming symptomatic or becoming infected given contact with an infectious individual.

The length of the the infectious and recovery period for each individual was set using a draw from a Poisson distribution:

$$f(x;\lambda) = \frac{e^{-\lambda}\lambda^x}{x!}$$

for x=0,1,2, where $\lambda$ is the average number of days until becoming infectious or the average number of days until recovering from infection.

We assumed the incubation and latency periods overlapped such that the start of symptoms (for those who were symptomatic) coincided with the beginning of the infectious period. Both the average length of the incubation and the recovery periods were not influenced by whether individuals were symptomatic (vs asymptomatic) or vaccinated. As a simplification, we also assumed that asymptomatic and symptomatic individuals were equally infectious. Besides vaccination, the population had a homogeneous risk for SARS-Cov-2.

COVID-like Symptoms

We assumed a constant proportion of individuals had COVID-like symptoms (not due to SARS-Cov-2). Individuals were assigned to have COVID-like symptoms via a random draw such that when one individual "recovered", another individual would replace them to maintain a constant proportion of the population with COVID-like symptoms. The length of the the recovery period for each individual with COVID-like symptoms was set using a draw from a Poisson distribution:

$$f(x;\lambda) = \frac{e^{-\lambda}\lambda^x}{x!}$$

For x=0,1,2, where $\lambda$ is the average number of days until recovering from infection. Note that individuals could also be simultaneously infected with SARS-CoV-2. Individuals could also have multiple occurrences of COVID-like symptoms from other etiologies over time.

Testing and Healthcare Engagement

Testing for SARS-CoV-2 was set to be a cumulative probability of 0.85 and 0.15 for individuals with high and low healthcare engagement across 10 days, respectively. We selected a 10 day period to reflect the average time individuals are symptomatic and infected with SARS-CoV-2 or have COVID-like symptoms. We gave each symptomatic individual an opportunity to test on each day. To convert our cumulative probability into a daily probability, we used the following formula $1-(1-p)^{10}$ where solving for p gives us the daily probability of failure (i.e. testing) over 10 days (assuming the probability of testing each day is independent). For simplicity, we assumed only symptomatic individuals were tested, that the etiologies of symptoms did not influence testing, and that SARS-CoV-2 tests had perfect sensitivity and specificity.

Testing Scenarios

We simulated the three testing scenarios by varying the degree of testing by vaccination status (“c” in Fig. 1). As the value of (“c”) is shaped by the relationships between healthcare engagement with vaccination (pathway marked “a” in Fig. 1) and with testing (pathway marked as “b” in Fig. 1), we varied (“c”) by modifying the relationship between the likelihood of vaccination and the level of healthcare engagement (“a”). Specifically, we assumed the vaccinated population had a constant proportion of individuals with high healthcare engagement (55%; based on a survey on testing intentions by vaccination status [5]) across the scenarios and then varied the proportion of high healthcare engagement in the unvaccinated population (scenario 1: 0.55 [equal vaccinated and unvaccinated], scenario 2: 0.22 [vaccinated 2.5x higher than unvaccinated] and scenario 3: 0.11 [vaccinated 5x higher than unvaccinated]). The proportion of low healthcare engagement for both groups was calculated as 1 - proportion of high healthcare engagement. Scenario 2 represents a relationship between vaccinated and unvaccinated found in empirical survey results[5]. Across all testing scenarios, the relationship between the likelihood of testing by the level of healthcare engagement (“b”) was kept constant (the probabilities of testing for high and low-healthcare engagement were set to be 0.85 and 0.15, respectively).

For each testing scenario, to determine the degree of testing difference by vaccination status (the strength of “c” in Fig. 1), we calculated the ratio of the testing probabilities by vaccination status:

$$Testing.Diff = \frac{T_V}{T_U}$$

where $T_V$ is the average probability of testing given you are vaccinated and $T_U$ is the average probability of testing given you are unvaccinated. Average testing probabilities $T_i$ where (i $\epsilon$ V, U), were calculated with a weighted average using the proportion of individuals with low and high healthcare engagement by vaccination status and the cumulative probabilities of testing by level of healthcare engagement:

$$T_i = prop.HH_i * prob.test.HH + (1 - Prop.HH_i) * prob.test.LH$$

Where $prop.HH_i$ is the proportion of high healthcare engagement in population i, $prob.test.HH$ is the cumulative probability of testing after 10 days given an individual has high healthcare engagement; and $prob.test.LH$ is the cumulative probability of testing after 10 days given individuals have low healthcare engagement.

For all testing scenarios, $T_V = 0.535 = 0.55 * 0.85 + (1 - 0.55) * 0.15$

$T_U$ varies across the three testing scenarios as the proportion of high healthcare engagement varies:

Scenario 1: $T_U = 0.535 = 0.55 * 0.85 + (1 - 0.55) * 0.15$

Scenario 2: $T_U = 0.304 = 0.22 * 0.85 + (1 - 0.22) * 0.15$

Scenario 3: $T_U = 0.227 = 0.11 * 0.85 + (1 - 0.11) * 0.15$

The testing differences by vaccination status for each scenario are as follows:

Scenario 1: $Test.Diff = 1.00 = \frac{0.535}{0.535}$

Scenario 2: $Test.Diff = 1.76 = \frac{0.535}{0.304}$

Scenario 3: $Test.Diff = 2.36 = \frac{0.535}{0.227}$

Hence, across the three testing scenarios: in equal testing (scenario 1), unvaccinated and vaccinated have equal testing; in moderately unequal testing (scenario 2), vaccinated have 1.76x higher testing; and in highly unequal testing (scenario 3); vaccinated have 2.36x higher testing.

Event Scheduling

At the start of each epidemic realization, 10 individuals were randomly selected to be classified as infectious. At each time step, variables were updated by a series of discrete-time processes. The order of the events at each timestep were as follows:

1. Reduce the number of days left by one for those exposed to SARS-CoV-2
2. Reduce the number of days left by one for those infectious with SARS-CoV-2
3. Reduce the number of days left by one for those with other etiologies of COVID-like symptoms
4. Change status from SARS-CoV-2 Exposed to SARS-CoV-2 Infectious if there were zero days remaining in SARS-CoV-2 exposure time
    a. Set each individual's symptom status to either Asymptomatic or Symptomatic using a random draw from a distribution
5. Change status from SARS-CoV-2 Infectious to Recovered if there were zero days remaining in SARS-CoV-2 infectious time; change other etiologies of COVID-like symptoms status to Non-infected if there were zero days remaining in other etiologies infection time.
6. Infect individuals with SARS-CoV-2 and set the SARS-CoV-2 status to Exposed
7. Infect individuals with other etiologies of COVID-like symptoms and set the status of other etiologies to Infected
    a. Set each individual's symptom status as symptomatic
8. Infected individuals with SARS-CoV-2 and symptomatic engage in a probability of testing given their level of health-seeking
    a. Testing status for SARS-CoV-2 is set to either Tested or remains as Non-tested

9. Infected individuals with other etiologies of COVID-like symptoms (who are automatically symptomatic) engage in a probability of testing given their level of healthcare engagement.
    a. Testing status for other etiologies of COVID-like symptoms set to either Tested or remains as Non-tested

Model Verification

For the cohort design, we calculate symptomatic VE using our simulated cohort and formula 1:

(1) $$VE_{RR} = 1 - RR(t)$$

with:

$$RR(t) = \frac{\frac{CI_V(t)}{N_V}}{\frac{CI_U(t)}{N_U}}$$

where $RR(t)$ is a relative risk at time *t*; $CI_V\ (t)$ and $CI_U\ (t)$ are the cumulative numbers of symptomatic infected for vaccinated and unvaccinated that have been tested and diagnosed (tested positive) at time *t,* respectively; and $N_V$ and $N_U$ are the total numbers of vaccinated and unvaccinated individuals, respectively.

For the test-negative design, we calculate VE using individuals from our simulated cohort who are both symptomatic and tested using formula 2:

(2) $$VE_{OR} = 1 - OR(t)$$

and

$$OR(t) = \frac{\frac{CI_V(t)}{CU_V(t)}}{\frac{CI_U(t)}{CU_U(t)}}$$

Where $OR\ (t)$ is an odds ratio at time *t*; and $CU_V\ (t)$ and $CU_U\ (t)$ are the cumulative numbers of symptomatic individuals (with COVID-like symptoms from other etiologies) for vaccinated and unvaccinated populations who tested negative for SARS-COV-2 and had no prior positive tests for SARS-COV-2 by time *t,* respectively.

To ensure our model was working correctly we compared estimates from our equal testing scenario with target estimates from the cohort design ($VE_{RR}$) and the test-negative design ($VE_{OR}$). We calculated $VE_{RR}$ and $VE_{OR}$ assuming everyone in the population who is eligible per study design criteria tested and then compared those outputs to our estimates from our equal testing scenario. We found that $VE_{RR}$ and $VE_{OR}$ estimates from the equal testing scenario results closely matched the target measurements of $VE_{RR}$ and $VE_{OR}$where everyone eligible per study design criteria had tested across both high and low levels of vaccine efficacy against susceptibility (high=0.55, low=0.1; Supplementary Fig. 15; Supplementary Fig. 16).

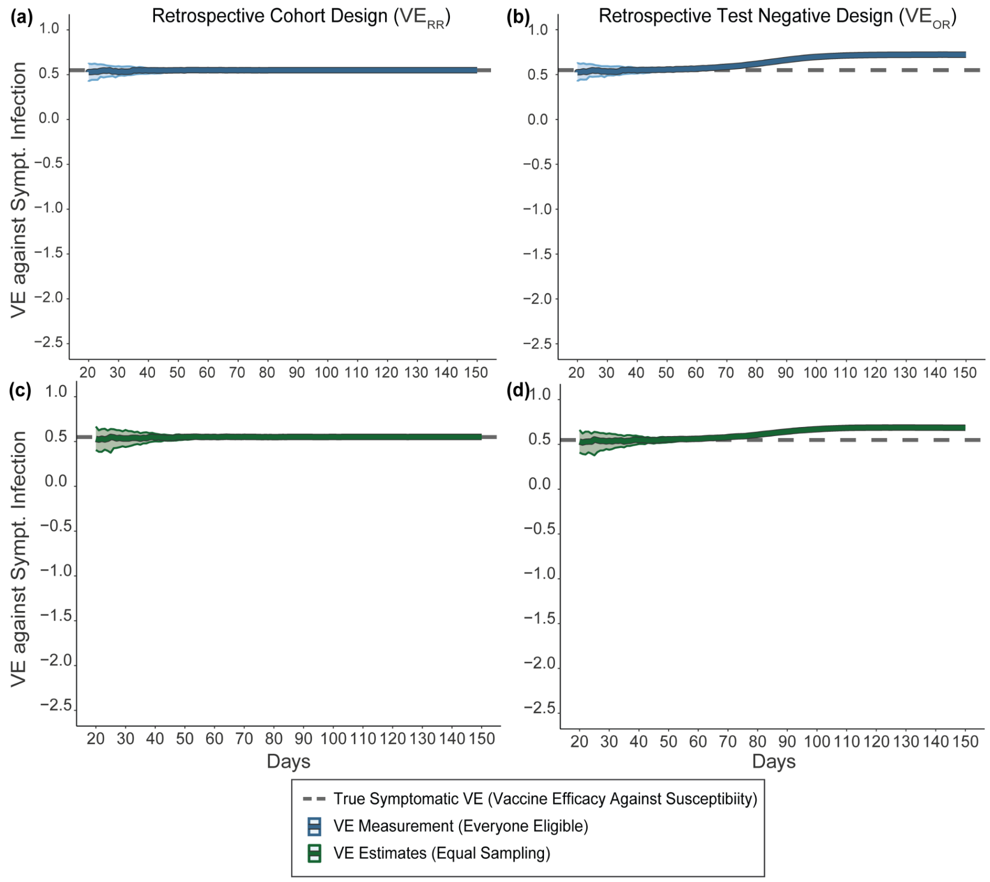


**Supplementary Fig. 15:** Symptomatic vaccine effectiveness measurements and estimates from the cohort design ( $VE_{RR}$;a and c) and the test-negative design ($VE_{OR}$; b and d) when vaccine efficacy against susceptibility is high (0.55). The true symptomatic VE (i.e. the level of vaccine efficacy against susceptibility) is depicted by the grey dashed line. VE measurements are the target measurements and include everyone who is eligible per study design criteria (dark blue line) while VE estimates are based on equal testing by vaccination status (dark green line). Central lines depict the median across 100 epidemic realizations, and the shaded area represents the interquartile range.

**Supplementary Fig. 16:** Symptomatic vaccine effectiveness measurements and estimates

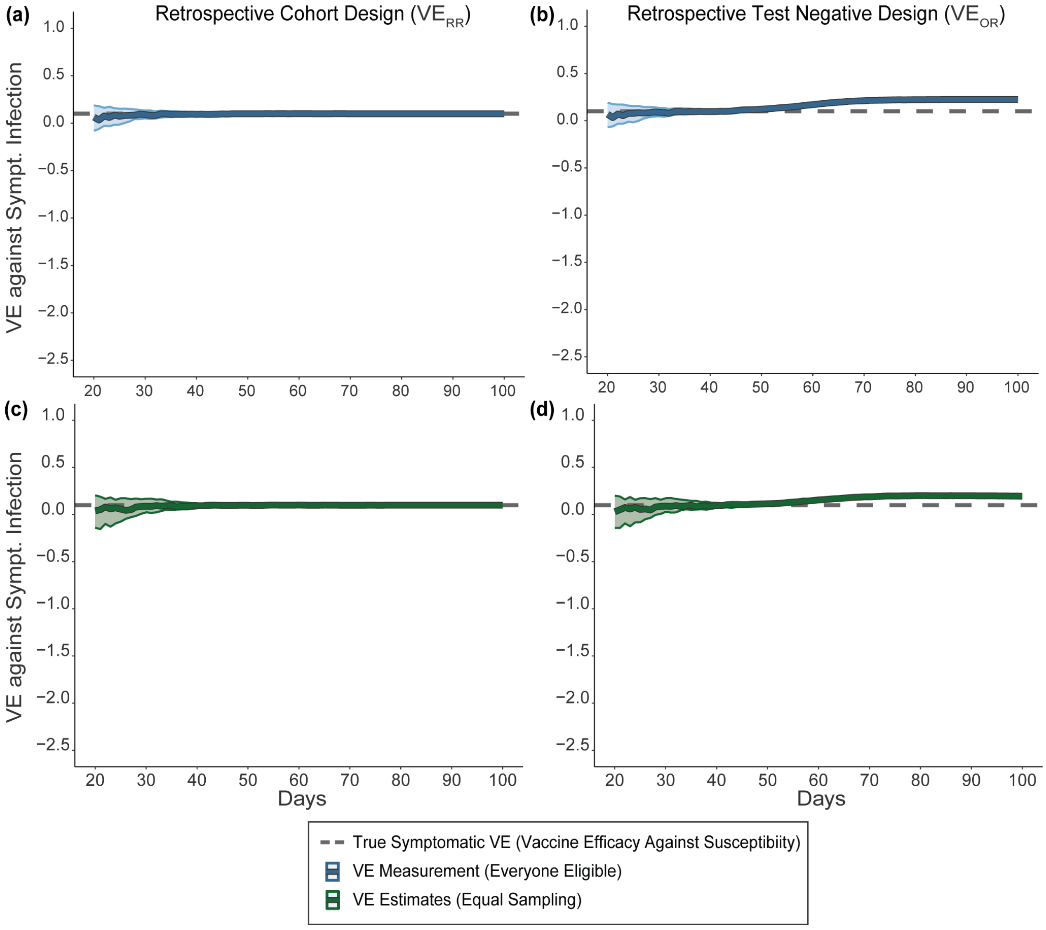


from the cohort design ($VE_{RR}$; a and c) and the test-negative design ($VE_{OR}$; b and d) when vaccine efficacy against susceptibility is low (0.1). The true symptomatic VE (i.e. the level of vaccine efficacy against susceptibility) is depicted by the grey dashed line. VE measurements are the target measurements and include everyone who is eligible per study design criteria (dark blue line) while VE estimates are based on equal testing by vaccination (dark green line). Central lines depict the median across 100 epidemic realizations and the shaded area represents the interquartile range.

Computing Resources

All simulations were conducted using R[6] (version 4.3.1) with computations performed on the Niagara supercomputer at the SciNet HPC Consortium[7,8]. Generating the random network for SARS-CoV-2 transmission and updating individual attributes during simulations and aggregating individual-level data for analyses was implemented using the R packages igraph[9] (version 1.5.1), graph4lg[10] (version 1.8.0) and data.table[11] (version 1.14.8). The code to replicate all analyses can be found on GitHub (https://github.com/mishra-lab/testing_bias.git).
